# Anxiety disorders and age-related changes in physiology

**DOI:** 10.1101/2021.08.15.21262059

**Authors:** Julian Mutz, Thole H. Hoppen, Chiara Fabbri, Cathryn M. Lewis

**Affiliations:** Social, Genetic and Developmental Psychiatry Centre, Institute of Psychiatry, Psychology & Neuroscience, King’s College London, London, United Kingdom; Institute of Psychology, University of Münster, Münster, Germany; Department of Biomedical and Neuromotor Sciences, University of Bologna, Bologna, Italy; Department of Medical and Molecular Genetics, Faculty of Life Sciences & Medicine, King’s College London, London, United Kingdom

**Keywords:** Ageing, Anxiety, Body composition, Cardiovascular function, Physiology

## Abstract

**Background:** Anxiety disorders are leading contributors to the global disease burden, highly prevalent across the lifespan, and associated with substantially increased morbidity and early mortality.

**Aims:** The aim of this study was to examine age-related changes across a wide range of physiological measures in middle-aged and older adults with a lifetime history of anxiety disorders compared to healthy controls.

**Method:** The UK Biobank study recruited >500,000 adults, aged 37–73, between 2006–2010. We used generalised additive models to estimate non-linear associations between age and hand-grip strength, cardiovascular function, body composition, lung function and heel bone mineral density in cases and in controls.

**Results:** The main dataset included 332,078 adults (mean age = 56.37 years; 52.65% females). In both sexes, individuals with anxiety disorders had a lower hand-grip strength and blood pressure, while their pulse rate and body composition measures were higher than in healthy controls. Case-control differences were larger when considering individuals with chronic and/or severe anxiety disorders, and differences in body composition were modulated by depression comorbidity status. Differences in age-related physiological changes between female anxiety disorder cases and healthy controls were most evident for blood pressure, pulse rate and body composition, while in males for hand-grip strength, blood pressure and body composition. Most differences in physiological measures between cases and controls decreased with increasing age.

**Conclusions:** Individuals with a lifetime history of anxiety disorders differed from healthy controls across multiple physiological measures, with some evidence of case-control differences by age. The differences observed varied by chronicity/severity and depression comorbidity.

## Introduction

Anxiety disorders are leading contributors to the global disease burden, highly prevalent across the lifespan and across nations, and associated with substantially increased morbidity and early mortality(1-4). A population-based study from Denmark reported that individuals with anxiety disorders had a 39% higher risk of premature death than the general population(5). This excess mortality does not result only from unnatural causes of death such as suicide, but also from increased rates of dementia, cardiovascular disease and other illnesses. Anxiety disorders are also associated with accelerated biological ageing, including earlier neurodegeneration(6, 7) and telomere attrition(8, 9), and an increased risk of disability in old age, especially in individuals with comorbid depression(10, 11). Less is known about physiological differences between individuals with anxiety disorders and healthy controls, and whether such differences vary by age. A Dutch longitudinal study reported that individuals with anxiety disorders had poorer lung function than healthy controls, and that men with anxiety disorders showed a greater decline in lung function over time(12). Women with anxiety disorders also had a lower hand-grip strength. More severe anxiety disorders were associated with greater physiological abnormalities. Most studies of physiology in anxiety disorders have focussed on one or two physiological measures(12-14) and research examining a range of physiological measures is lacking. To the best of our knowledge, this is the first study to examine age-related changes across a wide range of physiological measures in middle-aged and older adults with anxiety disorders. Most physiological measures can be assessed non-invasively, fast and at low cost, while providing reliable information on functional decline. Importantly, variation in physiological functioning predicts morbidity and mortality. A greater understanding of age-related physiological changes in anxiety disorders may inform strategies for prevention and intervention to foster healthy ageing.

### Aims

The aim of this cross-sectional study was to examine associations between age and 15 physiological markers in individuals with a lifetime history of anxiety disorders compared to healthy controls. Since the epidemiology of anxiety disorders(2) and human physiology(15) differ by sex, we conducted separate analyses in males and females. Given that a dose-response relationship between anxiety disorder severity and differences in physiology has been reported before(12), we also examined chronic and/or severe anxiety disorders. Finally, depression has been associated with age-related changes in physiology(16) and is highly comorbid with anxiety disorders(2-4, 12), hence we also examined individuals with anxiety disorders without comorbid depression.

## Method

### Study population

The UK Biobank is a prospective study of >500,000 adults aged 37–73 at baseline, recruited between 2006–2010. The study rationale and design have been described elsewhere(17). Briefly, individuals registered with the UK National Health Service (NHS) and living within a ∼40 km radius of one of 22 assessment centres were invited to participate. Participants provided information on their sociodemographic characteristics, lifestyle and medical history and underwent physical examination. Hospital inpatient records are available for most participants and primary care records are currently available for half of participants. A subset of 157,366 out of 339,092 invited participants (46%) completed an online follow-up mental health questionnaire (MHQ) between 2016 and 2017, covering 31% of all participants.

### Exposures

Age at the baseline assessment was the primary explanatory variable.

Details of the definition of cases and healthy controls are presented in Supplement 1. Briefly, we used a transdiagnostic phenotype for lifetime anxiety disorders and identified cases from multiple sources: the generalised anxiety disorder module of the Composite International Diagnostic Interview Short Form (CIDI-SF) (Supplement 2)(18) and the Generalised Anxiety Disorder Assessment (GAD-7)(19), which were assessed through the MHQ; psychiatric diagnoses reported during the nurse-led interview at baseline or in the MHQ; hospital inpatient records (Supplement 3); primary care records(20) (Supplement 4). We excluded individuals with bipolar disorder or psychosis, as these disorders are strongly associated with the risk of physical multimorbidity(21, 22). Healthy controls had no anxiety disorders, reported no current psychotropic medication use at baseline (Supplement 5)(23) and had no other mental disorders: no psychiatric diagnosis according to the nurse-led interview, MHQ, hospital inpatient or primary care records; no probable mood disorder(24) (Supplement 6); no Patient Health Questionnaire-9 (PHQ-9) sum score of ≥5; no GAD-7 sum score of ≥5; did not report ever feeling worried, tense, or anxious for most of a month or longer; no depression or bipolar disorder based on the CIDI-SF depression module and questions on (hypo)manic symptoms(16, 25).

### Physiological measures

We examined 15 physiological measures obtained at the baseline assessment, including maximal hand-grip strength, systolic and diastolic blood pressure, pulse rate, body mass index (BMI), waist-hip ratio, fat mass, fat-free mass, body fat percentage, peak expiratory flow, forced vital capacity (FVC), forced expiratory volume in one second (FEV_1_), FVC/FEV_1_ ratio, heel bone mineral density and arterial stiffness. Details of these measures have previously been reported in our study on age-related physiological changes in depression(16) and are presented in Supplement 7.

### Exclusion criteria

Participants whose genetic and self-reported sex did not match and participants with missing data, including “do not know” or “prefer not to answer”, for any covariates were excluded.

### Covariates

Covariates were identified from previous studies and included ethnicity, highest educational/professional qualification(26), physical activity (walking, moderate and vigorous-intensity activity), smoking status, alcohol intake frequency, sleep duration and, for cardiovascular measures, antihypertensive medication use (fields 6153 and 6177). Details of the sociodemographic and lifestyle factors are available in our previous publication(27).

### Statistical analyses

Analyses were prespecified prior to inspection of the data (preregistration: osf.io/hqu2f) and algorithms were tested on simulated data. Statistical analyses were conducted using R (version 3.6.0).

Sample characteristics were summarised using means and standard deviations or counts and percentages. Case-control differences were estimated using standardised mean differences (± 95% confidence intervals).

We examined the relationship between each physiological measure and age using generalised additive models (GAMs) with the ‘mgcv’ package in R(28). GAMs are flexible modelling approaches that allow for the relationship between an outcome variable and a continuous exposure to be represented by a non-linear smooth curve while adjusting for covariates. This approach is useful if a linear model does not capture key aspects of the relationship between variables and attempts to achieve maximum goodness-of-fit while maintaining parsimony of the fitted curve to minimize overfitting. Smoothing parameters were selected using the restricted maximum likelihood method and we used the default option of ten basis functions to represent smooth terms. Each measure was modelled against a penalised regression spline function of age with separate smooths for anxiety disorder cases and healthy controls.

Two models were fitted for each physiological measure in males and females separately:

- Unadjusted model: physiological measure ∼ anxiety disorder + s(age, by anxiety disorder).
- Adjusted model: physiological measure ∼ anxiety disorder + s(age, by anxiety disorder) + covariates (see previous paragraph).

where *s(age, by anxiety disorder)* represents the smooth function for age, stratified by anxiety disorder status.

To formally test whether the relationships between physiological measures and age differed between anxiety disorder cases and controls, we also fitted models that included reference smooths for healthy controls and difference smooths for anxiety disorder cases compared to healthy controls. For these analyses, anxiety disorder status was coded as an ordered factor in R. If the difference smooth differs from zero, the physiological measure follows a different trend with age in anxiety disorder cases and healthy controls.

Adjusted *p*-values were calculated using the *p*.*adjust* function in R to account for multiple testing across each set of analyses of the 15 physiological measures. Two methods were used: (1) Bonferroni and (2) Benjamini & Hochberg(29), two-tailed with *α* = .05 and false discovery rate of 5%, respectively. We have opted for this approach because the standard Bonferroni correction is usually too conservative, potentially leading to a high number of false negatives.

In a secondary analysis, we examined individuals with chronic and/or severe anxiety disorders, defined as individuals with (i) a hospital inpatient record of anxiety disorders as the primary diagnosis, (ii) recurrent or chronic anxiety (E2004 or E2005) in their primary care record or (iii) generalised anxiety disorder according to the CIDI-SF with maximum level of impairment (“Impact on normal roles during worst period of anxiety” (field 20418) = A lot) and duration (“Longest period spent worried or anxious” (field 20420) = All my life / as long as I can remember or at least 24 months).

In a sensitivity analysis, we excluded individuals with depression comorbidity from anxiety disorder cases(16).

We conducted two additional sensitivity analyses that were not pre-registered: (i) we additionally adjusted analyses of cardiovascular measures for BMI and (ii) we excluded anxiety disorder cases who reported current use of antidepressants at baseline from the analyses of blood pressure.

## Results

### Study population

A subset of 444,690 (88.49%) participants had complete data on all covariates. After excluding participants with missing physiological data, unclear anxiety disorder status (*n* = 93) or not meeting our inclusion criteria, we retained 332,078 participants in the main dataset. Subsets of 123,597, 228,321 and 107,958 participants were included in the analyses of lung function, heel bone mineral density and arterial stiffness, respectively (Supplement Figure 1).

### Sample characteristics

The average participant age in our main dataset was 56.37 years (SD = 8.11) and 52.65% of participants were female. Overall, 44,722 (13.47%) participants in this sample had a lifetime history of anxiety disorders, 65.92% (*n* = 29,482) of whom were female. Descriptive statistics for the full UK Biobank and for the analytical samples stratified by sex and anxiety disorder status are presented in Table 1 (physiological measures) and Supplement Table 4 (covariates).

**Table 1.** Physiological measures

| | Overall<br>($N=502,521$) | | Female | | Male | |
| --- | --- | --- | --- | --- | --- | --- |
| | | | Healthy control<br>($N=145,364$) | Anxiety disorder<br>($N=29,482$) | Healthy control<br>($N=141,992$) | Anxiety disorder<br>($N=15,240$) |
|  | Mean (SD) | Missing | Mean (SD) | Mean (SD) | Mean (SD) | Mean (SD) |
| Hand-grip strength | 31.77 (11.34) | 3203 | 24.89 (6.48) | 24.68 (6.64) | 41.25 (9.11) | 40.53 (9.22) |
| Systolic blood pressure | 137.87 (18.65) | 1325 | 135.71 (19.32) | 133.23 (18.48) | 141.28 (17.42) | 139.63 (16.92) |
| Diastolic blood pressure | 82.26 (10.15) | 1323 | 80.70 (9.96) | 80.29 (9.91) | 84.22 (9.96) | 83.92 (9.93) |
| Pulse rate | 69.42 (11.26) | 1323 | 69.87 (10.40) | 70.33 (10.68) | 67.76 (11.60) | 68.66 (12.04) |
| Body mass index | 27.43 (4.80) | 3105 | 26.66 (4.85) | 26.89 (5.20) | 27.64 (4.02) | 27.76 (4.33) |
| Body fat percentage | 31.45 (8.55) | 10408 | 36.06 (6.74) | 36.40 (6.96) | 24.97 (5.63) | 25.27 (5.81) |
| Fat mass | 24.86 (9.57) | 10973 | 26.14 (9.50) | 26.83 (10.23) | 21.87 (7.88) | 22.37 (8.48) |
| Fat-free mass | 53.22 (11.50) | 10176 | 44.33 (4.84) | 44.62 (5.05) | 63.67 (7.62) | 63.80 (7.89) |
| Waist-hip ratio | 0.87 (0.09) | 2265 | 0.81 (0.07) | 0.82 (0.07) | 0.93 (0.06) | 0.94 (0.06) |
| | | | $N=50,729$ | $N=10,128$ | $N=56,695$ | $N=6,045$ |
| Peak expiratory flow | 407.82 (130.78) | 168566 | 352.52 (76.81) | 356.88 (77.84) | 510.07 (115.61) | 509.72 (115.63) |
| Forced expiratory volume 1s | 2.85 (0.78) | 149197 | 2.48 (0.53) | 2.51 (0.53) | 3.43 (0.74) | 3.44 (0.74) |
| Forced vital capacity | 3.78 (0.98) | 149197 | 3.25 (0.64) | 3.30 (0.65) | 4.57 (0.88) | 4.58 (0.89) |
| FEV<sub>1</sub> / FVC | 0.75 (0.07) | 149197 | 0.76 (0.06) | 0.76 (0.06) | 0.75 (0.07) | 0.75 (0.07) |
| | | | $N=101,279$ | $N=19,217$ | $N=97,749$ | $N=10,076$ |
| Heel bone mineral density | 0.54 (0.14) | 180831 | 0.52 (0.12) | 0.52 (0.12) | 0.58 (0.15) | 0.57 (0.15) |
| | | | $N=45,320$ | $N=10,630$ | $N=46,549$ | $N=5,459$ |
| Arterial stiffness | 9.34 (4.05) | 332721 | 8.72 (4.27) | 8.71 (3.33) | 9.94 (4.59) | 9.94 (2.92) |

Note: SD = standard deviation; FEV<sub>1</sub> = forced expiratory volume in one second; FVC = forced vital capacity.

### Case-control differences

Case-control differences by sex are presented in Table 2. Females with anxiety disorders had a lower hand-grip strength and blood pressure than healthy controls. Their pulse rate was elevated, and they had higher values for all body composition measures and most lung function measures than controls. We did not find evidence of differences in the FEV_1_/FVC ratio, heel bone mineral density or arterial stiffness. Male anxiety disorder cases had a lower hand-grip strength, blood pressure and heel bone mineral density than healthy controls. Their pulse rate and all body composition measures were higher than in controls, although the difference in fat-free mass did not survive multiple testing correction. We did not find evidence of differences in lung function or arterial stiffness in males. The largest case-control difference was observed for systolic blood pressure (SMD = −0.129, 95% CI −0.142 to −0.117, *p*_Bonf._ < 0.001 in females and SMD = −0.091, 95% CI −0.111 to −0.078, *p*_Bonf._ = 0.013 in males).

**Table 2.**
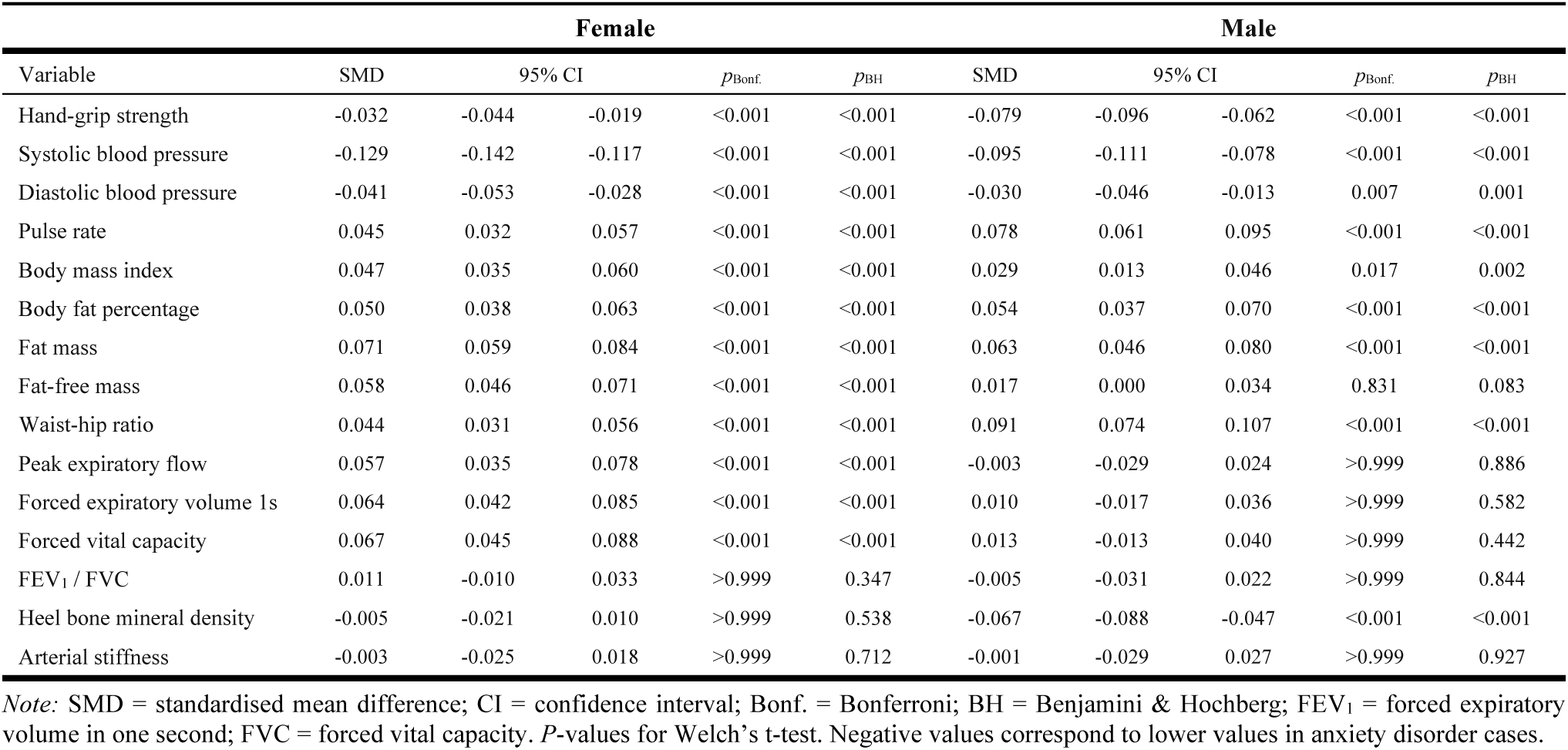
Differences in physiological measures between individuals with anxiety disorders and healthy controls

#### Chronic and/or severe anxiety

Between 8.37% to 9.46% of female and 9.47% to 9.99% of male cases had chronic and/or severe anxiety disorders (Supplement Table 5). In females, we observed the same overall pattern of results as in the main analysis, although all observed differences were larger in magnitude. For example, the case-control difference in body fat percentage was SMD = 0.113 (95% CI 0.074-0.151, *p*_Bonf._ < 0.001) (Supplement Table 6), compared to SMD = 0.050 (95% CI 0.038-0.063, *p*_Bonf._ < 0.001) in the main analysis. In male chronic and/or severe cases, we did not find evidence of a difference in diastolic blood pressure compared to healthy controls (SMD = −0.013, 95% CI −0.064 to 0.038, *p*_BH_ = 0.713) and the difference in body fat percentage was not statistically significant after multiple testing correction. For most other physiological measures, we observed larger case-control differences than in the main analysis (Supplement Table 6). For example, the case-control difference in systolic blood pressure was SMD = −0.095 (95% CI −0.111 to −0.078, *p*_Bonf._ < 0.001) in the main analysis and SMD = −0.153 (95% CI −0.204 to −0.102, *p*_Bonf._ < 0.001) in this analysis.

#### Anxiety without depression comorbidity

After excluding anxiety disorder cases with depression comorbidity, we retained between 25.91% to 37.67% of female and 31.45% to 44.69% of male cases (Supplementary Table 5). In females, differences in hand-grip strength, blood pressure, pulse rate and arterial stiffness remained statistically significant but were smaller in magnitude than in the main analysis (Supplement Table 7). Body mass index, fat mass and fat-free mass, which were higher in cases than in controls in the main analysis, were lower in cases without depression comorbidity (SMDs between −0.026 and −0.049). We did not find evidence of case-control differences in body fat percentage or waist-hip ratio in this analysis. There was also no longer evidence of differences in lung function, except that the FEV_1_/FVC ratio was lower in cases than in controls (SMD = −0.048, 95% CI −0.082 to −0.013, *p*_BH_ = 0.019). Finally, heel bone mineral density was lower in female cases (SMD = −0.036, 95% CI −0.060 to −0.012, *p*_Bonf._ = 0.043). In males, differences in hand-grip strength, blood pressure, pulse rate, waist-hip ratio, lung function, heel bone mineral density and arterial stiffness were similar to the main analysis. Body mass index and fat-free mass were lower in cases than in controls (SMD = −0.058, 95% CI −0.084 to −0.033, *p*_Bonf._ < 0.001 and SMD = −0.066, 95% CI −0.092 to −0.041, *p*_Bonf._ < 0.001, respectively) and we did not find evidence of case-control differences in body fat percentage or fat mass, which were elevated in cases in the main analysis.

### Case-control differences by age

We found some evidence that age-related changes in blood pressure, pulse rate, body composition and heel bone mineral density differed between female cases and controls (Figure 1). Systolic blood pressure was −0.9 mmHg lower in cases at age 45 and this difference widened to −2.2 mmHg at age 65. For diastolic blood pressure, we did not find evidence of case-control differences below age 52, and slightly lower diastolic blood pressure in cases than in controls above age 52. Case-control differences in pulse rate and body composition narrowed with age (Supplement Figure 2). Heel bone mineral density was slightly lower in cases than in controls below age 55, and there was no evidence of differences between older cases and controls.

**Figure 1.**
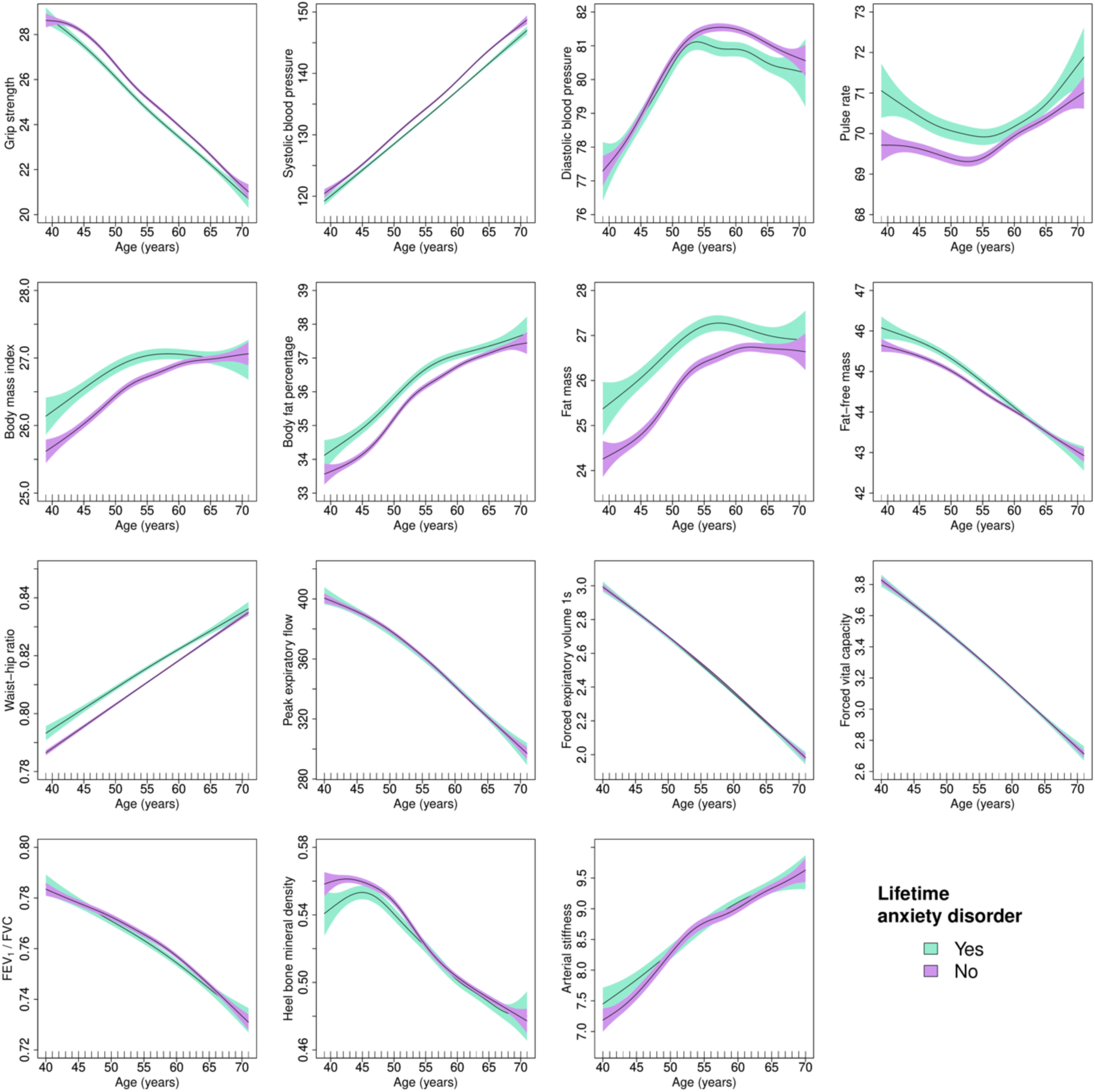
Generalised additive models of age-related changes in physiological measures in females with anxiety disorders and healthy controls. The solid lines represent physiological measures against smoothing functions of age. The shaded areas correspond to approximate 95% confidence intervals (± 2 × standard error). FEV1 = forced expiratory volume in one second; FVC = forced vital capacity.

In males, case-control differences in hand-grip strength, pulse rate, waist-hip ratio and heel bone mineral density narrowed with age (Figure 2 and Supplement Figure 3). There was some evidence that diastolic blood pressure was lower in cases than in controls above age 50, although the formal statistical test did not survive multiple testing correction. We found little evidence of case-control differences by age for the other physiological measures.

**Figure 2.**
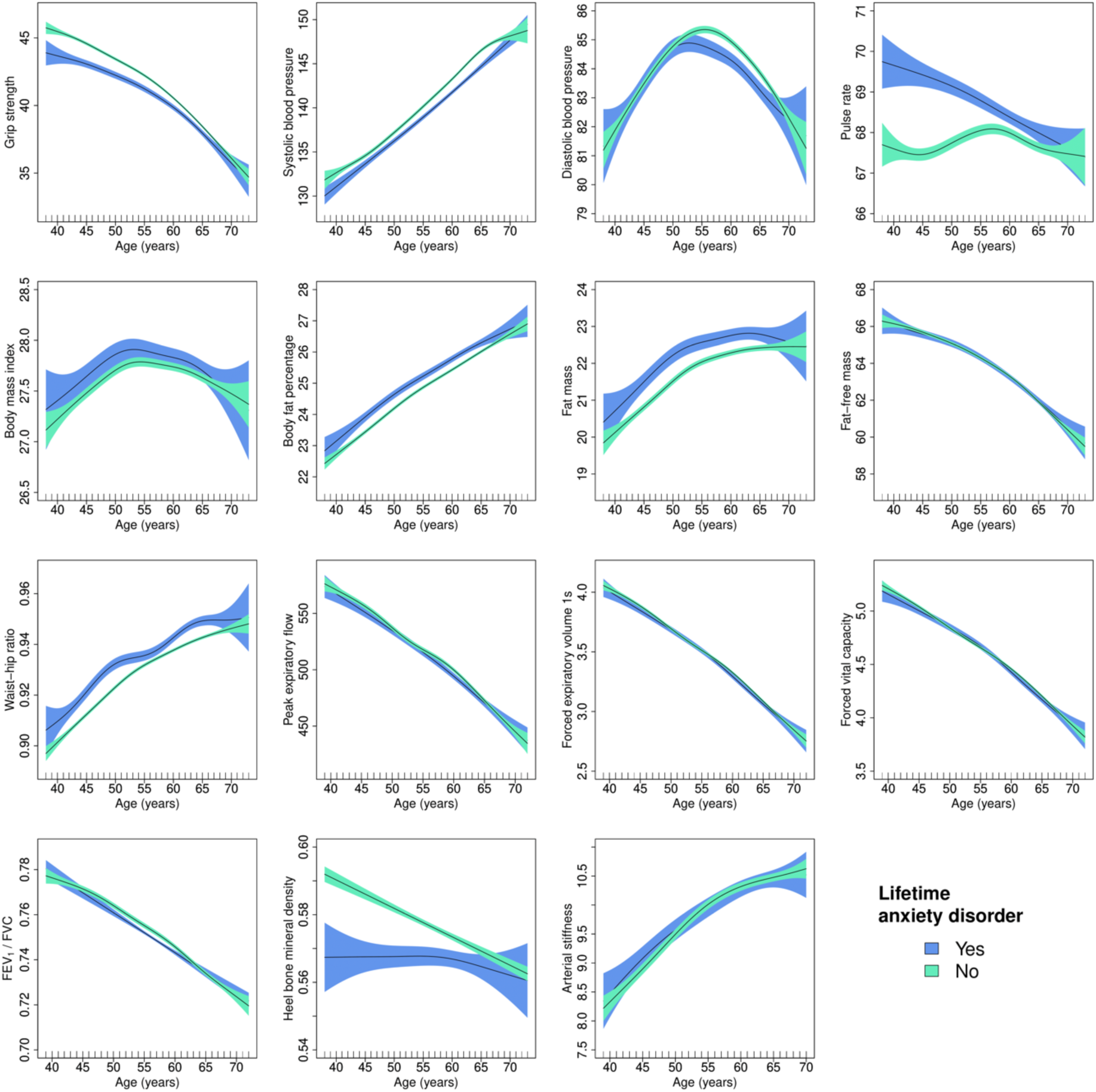
Generalised additive models of age-related changes in physiological measures in males with anxiety disorders and healthy controls. The solid lines represent physiological measures against smoothing functions of age. The shaded areas correspond to approximate 95% confidence intervals (± 2 × standard error). FEV_1_ = forced expiratory volume in one second; FVC = forced vital capacity.

In females, we observed similar results across all physiological measures after adjustment for covariates (Supplement Figures 4 and 5). The formal statistical tests provided no evidence of case-control differences in age-related changes in heel bone mineral density, however, the overall pattern of results was comparable to the unadjusted model. In males, we also observed similar results in the adjusted model (Supplement Figures 6 and 7). Although the trajectories were similar to the unadjusted analysis, the formal statistical tests provided no evidence of case-control differences in age-related changes in waist-hip ratio. The same was true for diastolic blood pressure and heel bone mineral density after multiple testing correction.

#### Chronic and/or severe anxiety

In female chronic and/or severe cases, we observed similar results for age-related changes in blood pressure and body composition, except that there was no evidence of case-control differences in fat-free mass below age 45 (Supplement Figures 8 and 9). As in the main analysis, we observed no evidence of case-control differences in age-related changes in lung function or arterial stiffness. Differences in results were most evident for hand-grip strength and pulse rate, and to a lesser extent for heel bone mineral density. The formal statistical tests provided some evidence of case-control differences in age-related changes in blood pressure, body mass index, fat mass and fat-free mass, although none survived multiple testing correction. For male chronic and/or severe cases, we found some evidence that case-control differences in hand-grip strength and pulse rate narrowed with age, similar to the results from the main analysis, although none of the formal statistical tests survived multiple testing correction. There was less evidence of case-control differences in age-related changes in systolic blood pressure and no evidence of case-control differences in age-related changes in diastolic blood pressure. For all other physiology measures, none of the formal statistical tests were statistically significant (Supplement Figures 10 and 11).

#### Anxiety without depression comorbidity

In female cases without depression comorbidity, none of the formal statistical tests provided evidence of case-control differences in age-related changes in physiology. We also observed less evidence of differences by age in blood pressure or pulse rate. There was some evidence that several body composition and lung function measures were lower in cases than in controls between ages 45 to 65 (Supplement Figures 12 and 13). In male cases, the formal statistical test provided some evidence of case-control differences in age-related changes in pulse rate, although it did not survive multiple testing correction. None of the other formal statistical tests were statistically significant and there was less evidence of case-control differences in age-related physiological changes (Supplement Figures 14 and 15).

### Additional sensitivity analyses

Case-control differences in cardiovascular function by age were similar to the results of our main analysis after additional adjustment for BMI (Supplement Figures 16 and 17). Excluding anxiety disorder cases who reported current use of antidepressants (*n* = 6517 females and *n* = 2700 males) had a negligible effect on case-control differences in blood pressure (Supplement Table 7).

## Discussion

### Principal findings

We observed case-control differences in hand-grip strength, blood pressure, pulse rate and body composition in both sexes, while case-control differences in lung function and heel bone mineral density were specific to females and males, respectively. We found no evidence of case-control differences in arterial stiffness.

Most of the observed differences were larger when we examined chronic and/or severe anxiety disorders. However, in males, the difference in diastolic blood pressure was smaller and not statistically significant, and differences in body fat percentage and heel bone mineral density, while similar in magnitude, were no longer statistically significant.

After excluding cases with comorbid depression, most differences remained statistically significant but were smaller in magnitude. However, body composition measures in both sexes were either lower in cases, or we did not find evidence of case-control differences. We found some evidence that heel bone mineral density was lower in female cases without comorbid depression. Most case-control differences in lung function, however, were no longer statistically significant. We also did not find evidence of case-control differences in diastolic blood pressure in males without comorbid depression.

Differences in age-related physiological changes between female anxiety disorder cases and healthy controls were most evident for blood pressure, pulse rate and body composition, with some evidence of differences in heel bone mineral density. Most case-control differences narrowed with age, except that we found a larger difference in blood pressure in older participants. In males, case-control differences in hand-grip strength, pulse rate, and to a lesser extent in waist-hip ratio and heel bone mineral density narrowed with age. Diastolic blood pressure was lower in older cases than in controls.

The overall pattern of results was comparable in female chronic and/or severe cases, but there was generally less evidence of differences between trajectories in males. Except for body composition, there was limited evidence of case-control differences in age-related physiological changes after excluding individuals with comorbid depression.

### Findings in context

Consistent with findings from the Netherland Study of Depression and Anxiety (NESDA)(12), female cases had a lower hand-grip strength. Hand-grip strength was also lower in male cases, which had not been observed in the NESDA.

Previous findings regarding anxiety disorders and blood pressure have been mixed and studies have often examined hypertension instead of blood pressure(30-33). While a recent meta-analysis reported increased rates of hypertension in anxiety disorders(14), several studies found no statistically significant associations with hypertension or blood pressure(34-38). Some studies observed lower blood pressure in anxiety disorders(39-41). Antidepressant medication and benzodiazepine use may affect blood pressure(42-44), and some population-based studies have observed lower blood pressure in depression(16, 42). The high degree of comorbidity between anxiety disorders and depression and differences in medication use might partially explain these mixed results. We observed lower blood pressure in anxiety disorders, except for diastolic blood pressure in males, irrespective of depressive comorbidity. Excluding individuals who reported antidepressant use at baseline resulted in a negligible decrease in the case-control difference in systolic blood pressure and a negligible increase in the difference in diastolic blood pressure. Case-control differences in blood pressure were larger in chronic and/or severe anxiety disorders.

Consistent with previous research(45, 46), we observed a higher pulse rate in anxiety disorder cases. Noteworthy, reductions in anxiety disorder severity following cognitive behavioural therapy have been associated with a decrease in resting pulse rate(47).

Previous research has found higher rates of obesity(48, 49) and poor diet(50) in anxiety disorders, consistent with our observation that cases had elevated measures of body composition. However, our analyses suggested that these differences may be modified by depression comorbidity. Depression has been associated with increased metabolic risk factors(51) and elevated body composition measures(16). One study found that depression, but not anxiety disorders, was associated with an increased risk of metabolic syndrome(52), although a large Finnish birth cohort study found no evidence that either depression or anxiety disorders were associated with metabolic syndrome(53).

Our lung function results contradict previous research. A cross-sectional analysis of the NESDA found poorer lung function in females with anxiety disorders and/or depression compared to healthy controls and better lung function in male cases than in controls(54). A 6-year longitudinal assessment of these participants suggested a greater decline in lung function in male cases compared to controls(12). We observed better lung function in female cases and found no statistically significant case-control differences in males. However, we observed no evidence of differences after excluding individuals with comorbid depression. Noteworthy, the results from the NESDA were not reported separately for anxiety disorders and depression.

Our results confirm previous research(55) that found a lower bone mineral density in male cases and limited evidence of a case-control difference in females.

We did not observe any differences in arterial stiffness between cases and controls. This finding is surprising given that previous studies have reported increased arterial stiffness in anxiety disorders(56, 57).

Inconsistencies in findings between studies could result from differences in sample characteristics such as age, sex, severity of symptoms or the prevalence of effect modifiers. Systematic reviews of studies examining each measure of physiological function in relation to anxiety disorders could address these important questions. The definition of anxiety disorders, including which specific diagnoses were included, might also contribute to differences in results.

To our knowledge, we report the first study of age-related changes in physiology in anxiety disorders with age as a continuous rather than a categorical variable. Previous studies dichotomised age, for example testing differences between middle-aged participants and others(14), making comparisons with our study difficult.

### Mechanisms

Several mechanisms could explain the physiological differences between anxiety disorders cases and healthy controls. Anxiety disorders are sometimes associated with less healthy lifestyle behaviours(58) which could affect a range of physiological makers. Physiological differences could also reflect the cumulative effects of anxiety-related overactivation of the hypothalamic-pituitary-adrenal axis and sympathetic nervous system(33) as well as increased inflammation and oxidative stress(59-61). It is also possible that the reciprocal relationship between late-life anxiety and associated cognitive impairment may be a driver of poor physiological function(62). The greater case-control differences observed in chronic and/or severe anxiety disorders could be explained by a dose-response relationship between severity of symptoms and physiology mediated by a greater impact of anxiety on lifestyle, overactivation of the sympathetic nervous system and other potential pathways. A potential explanation for the observation that case-control differences decreased with increasing age is that the prevalence of comorbidities increases with age which may dilute the effects that anxiety disorders may have.

### Limitations

The cross-sectional study design presents uncertainty about whether case-control differences by age represent changes due to ageing or potential cohort effects. Future work should examine physiological function in anxiety disorders longitudinally. Although we found that all physiological measures varied by age, selection bias resulting in healthier older adults participating at higher rates relative to their age group could result in the underestimation of age-related changes. To achieve maximum cohort coverage, we identified cases from multiple data sources, with strengths and limitations that have been discussed elsewhere(23, 63, 64). For the primary care records, we included the additional quality control criterion that cases needed to have at least two mentions of anxiety in their records. Anxiety disorder cases in this study also included individuals with a single episode. This likely resulted in an underestimation of case-control differences. A small number of individuals with subthreshold disorders could be present amongst the healthy controls, which could have attenuated observed differences. We examined a transdiagnostic definition of anxiety disorders, and differences in physiological function between specific diagnoses could be explored in future studies. Although we found that the differences between individuals with chronic and/or severe anxiety disorders and controls were larger, these differences could at least partly reflect higher levels of comorbidities in addition to the effects of chronicity and/or severity. Some caution is warranted in interpreting the findings of the sensitivity analyses due to the smaller sample size and lower statistical power. Finally, limitations in the assessment of physiological function may have masked some differences between cases and controls. For example, dynamic measures such as pulse rate were not measured longitudinally, and our study does therefore not provide insights into potential case-control differences that could be identified from time-series data. Similarly, lung function was estimated through volumetric measures using breath spirometry and we could therefore not examine irregularity of respiratory patterns. Physiological measures with higher temporal resolution could be examined in future studies.

### Generalisability

UK Biobank participants are not fully representative of the UK population. MHQ respondents were also more educated, of higher socioeconomic status and had fewer long-standing illnesses than participants who did not complete the MHQ. Similar patterns of disease prevalence were present in the MHQ and hospital inpatient records, although anxiety was reported more frequently in the MHQ(18). Nevertheless, the overall number of individuals with a lifetime history of anxiety disorders identified in our study was comparable to previous epidemiological studies(65). Wider issues of generalisability of findings from the UK Biobank have been discussed elsewhere(66). Our findings do not generalise to populations below age 40 or older than age 70, and there was greater uncertainty near the lower and upper extremes of the age range in this study. Additional studies in younger participants and in the elderly are needed.

### Implications

Individuals with a lifetime history of anxiety disorders differed from healthy controls across multiple physiological measures, with some evidence of case-control differences by age. The differences observed varied by chronicity/severity and depression comorbidity. Monitoring of physiological function in individuals with anxiety disorders should be adapted depending on depression comorbidity status.

### Ethics

We assert that all procedures contributing to this work comply with the ethical standards of the relevant national and institutional committees on human experimentation. All procedures were approved for the UK Biobank study by the National Information Governance Board for Health and Social Care and the NHS North West Multicentre Research Ethics Committee (11/NW/0382). No project-specific ethical approval is needed. Data access permission has been granted under UK Biobank application 45514. Written informed consent was obtained from all participants.

## Data Availability

The data used are available to all bona fide researchers for health-related research that is in the public interest, subject to an application process and approval criteria. Study materials are publicly available online at http://www.ukbiobank.ac.uk.

## Acknowledgments

This research has been conducted using data from UK Biobank, a major biomedical database. This project made use of time on Rosalind HPC, funded by Guy’s & St Thomas’ Hospital NHS Trust Biomedical Research Centre (GSTT-BRC), South London & Maudsley NHS Trust Biomedical Research Centre (SLAM-BRC), and Faculty of Natural Mathematics & Science (NMS) at King’s College London.

## Author Contribution

CML acquired the studentship funding, interpreted the findings and critically reviewed the manuscript. JM conceived the idea of the study, acquired the data, carried out the statistical analysis, interpreted the findings, wrote the manuscript and revised the manuscript for final submission. THH contributed to the study design, interpreted the findings and contributed to the writing of the manuscript. CF interpreted the findings and critically reviewed the manuscript. All authors read and approved the final manuscript. JM had full access to all data used in this study and takes responsibility for the integrity of the data and the accuracy of the data analysis.

