## Supplement for "Anxiety disorders and age-related changes in physiology"

The following material accompanies the article

**Supplement 1. Case control criteria**

We used a transdiagnostic phenotype for lifetime anxiety disorders and identified cases from multiple sources: the MHQ which included the anxiety disorder module of the Composite International Diagnostic Interview Short Form (CIDI-SF) and assessed generalised anxiety disorder according to DSM-5 criteria (Supplement 1)[1]; individuals with a Generalised Anxiety Disorder Assessment (GAD-7) sum score of ≥10[2], which was assessed as part of the MHQ; individuals who had reported "anxiety/panic attacks" during the nurse-led interview at baseline (field 20002), or "anxiety, nerves or generalized anxiety disorder", "social anxiety or social phobia", "any other phobia", "panic attacks" or "agoraphobia" in response to a single-item question on the MHQ (field 20544); participants with a hospital inpatient record containing an ICD-10 code for anxiety disorders (F40-F41; Supplement 2); participants with at least two primary care records containing a Read v2 or CTV3 code for anxiety disorders (for data extraction procedures, see [3]) (Supplement 3). We excluded individuals with any record of bipolar disorder or psychosis, as these disorders are strongly associated with the risk of physical multimorbidity[4,5].

Healthy controls did not meet our criteria for anxiety disorders and had no record of other mental disorders: (i) had not reported "schizophrenia", "mania/bipolar disorder/manic depression", "depression", "obsessive compulsive disorder", "anorexia/bulimia/other eating disorder", "post-traumatic stress disorder" during the nurse-led interview at baseline (field 20002); (ii) reported no mental disorders in response to the single-item question on the MHQ (field 20544); (iii) had self-reported no current psychotropic medication use at baseline (field 20003; Supplement 4)[6]; (iv) had no linked hospital inpatient record that contained any ICD-10 Chapter V code except organic causes or substance use (F20-F99); (v) had no primary care record containing diagnostic codes for mental disorders[3]; (vi) were not classified as individuals with probable mood disorder according to Smith et al.[7] based on additional questions that were introduced during the later stages of the baseline assessment (Supplement 5); (vii) had no Patient Health Questionnaire-9 (PHQ-9) sum score of ≥5, which was assessed as part of the MHQ; (viii) had no GAD-7 sum score of ≥5; (ix) did not report that they ever felt worried, tense, or anxious for most of a month or longer (field 20421); (x) were not identified as depression or bipolar disorder cases based on the CIDI-SF depression module and questions on (hypo)manic symptoms[8,9].

### Supplement 2. CIDI-SF lifetime generalised anxiety disorder criteria

**Supplement table 1.** Case definition lifetime generalised anxiety disorder

| | Definition and UK Biobank data fields |
|---|---|
| **Lifetime generalised anxiety disorder** (case):<br>Excessive anxiety or worry about a number of issues, occurring most days for at least six months, finding it difficult to control the worry, with three or more somatic symptoms, and functional impairment.<br><br>No record of psychosis or bipolar disorder. | "Ever felt worried, tense, or anxious for most of a month or longer" (20421) = Yes<br>AND<br>"Longest period spent worried or anxious" (20420) ≥ 6 months or All my life<br>AND<br>"Worried most days during period of worst anxiety" (20538) = Yes<br>AND<br>("Ever worried more than most people would in similar situation" (20425) = Yes OR "Stronger worrying (than other people) during period of worst anxiety" (20542) = Yes)<br>AND<br>("Number of things worried about during worst period of anxiety" (20543) = More than one thing OR "Multiple worries during worst period of anxiety" (20540) = Yes)<br>AND<br>("Difficulty stopping worrying during worst period of anxiety" (20541) = Yes OR "Frequency of inability to stop worrying during worst period of anxiety" (20539) = Often OR "Frequency of difficulty controlling worry during worst period of anxiety" (20537) = Often)<br>AND<br>"Impact on normal roles during worst period of anxiety" (20418) = Somewhat or A lot<br>AND<br>Three or more somatic symptoms endorsed from: (1) "Restless during period of worst anxiety" (20426), (2) "Keyed up or on edge during worst period of anxiety" (20423), (3) "Easily tired during worst period of anxiety" (20429), (4) "Difficulty concentrating during worst period of anxiety" (20419), (5) "More irritable than usual during worst period of anxiety" (20422), (6) "Tense, sore, or aching muscles during worst period of anxiety" (20417), (7) "Frequent trouble falling or staying asleep during worst period of anxiety" (20427).<br>AND<br>**No** self-reported psychosis or mania for "Mental health problems ever diagnosed by a professional" (20544)<br>AND<br>**No** self-reported mania/bipolar disorder/manic depression or schizophrenia for "Non-cancer illness" (20002)<br>AND<br>**No** ICD-10 code for "manic episode" or "bipolar affective disorder" (F30-F31) or "schizophrenia, schizotypal and delusional disorders" (F20-F29)<br>AND<br>**No** probable bipolar disorder (20126) according to Smith et al. (2013)<br>AND<br>**No** bipolar disorder record according to the MHQ<br>AND<br>**No** primary care record of bipolar disorder or psychosis |

*Note:* Criteria for lifetime generalised anxiety disorder adapted from Davis et al. (2020), doi: 10.1192/bjo.2019.100. CIDI-SF = Composite International Diagnostic Interview Short Form; ICD-10 = International Classification of Diseases, Tenth Revision; MHQ = mental health questionnaire.

**Supplement 3. ICD-10 codes for anxiety disorders**

**Phobic anxiety disorders**

- F40.0 Agoraphobia
- F40.1 Social phobias
- F40.2 Specific (isolated) phobias
- F40.8 Other phobic anxiety disorders
- F40.9 Phobic anxiety disorder, unspecified

**Other anxiety disorders**

- F41.0 Panic disorder [episodic paroxysmal anxiety]
- F41.1 Generalised anxiety disorder
- F41.2 Mixed anxiety and depressive disorder
- F41.3 Other mixed anxiety disorders
- F41.8 Other specified anxiety disorders
- F41.9 Anxiety disorder, unspecified

### Supplement 4. Primary care codes for anxiety disorders

**Supplement table 2.** Primary care record codes

| Clinical code | Diagnostic version | Code description |
|---|---|---|
| 146G. | Read_v3 | H/O: agoraphobia |
| 1Bb.. | Read_v2 | Specific fear |
| 1Bb0. | Read_v2 | Fear of falling |
| 1Bb1. | Read_v2 | Fear of getting cancer |
| E200. | Read_v3 | Anxiety disorder |
| E200. | Read_v2 | Anxiety states |
| E2000 | Read_v3 | Anxiety state unspecified |
| E2000 | Read_v2 | Anxiety state unspecified |
| E2001 | Read_v3 | (Panic disorder) or (panic attack) |
| E2001 | Read_v2 | Panic disorder |
| E2002 | Read_v3 | Generalised anxiety disorder |
| E2002 | Read_v2 | Generalised anxiety disorder |
| E2004 | Read_v3 | Chronic anxiety |
| E2004 | Read_v2 | Chronic anxiety |
| E2005 | Read_v3 | Recurrent anxiety |
| E2005 | Read_v2 | Recurrent anxiety |
| E200z | Read_v3 | Anxiety state NOS |
| E200z | Read_v2 | Anxiety state NOS |
| E202. | Read_v3 | Phobic disorders (& [social] or [phobic anxiety]) |
| E202. | Read_v2 | Phobic disorders |
| E2020 | Read_v3 | Phobia unspecified |
| E2020 | Read_v2 | Phobia unspecified |
| E2021 | Read_v3 | Agoraphobia with panic attacks |
| E2021 | Read_v2 | Agoraphobia with panic attacks |
| E2022 | Read_v3 | Agoraphobia without mention of panic attacks |
| E2022 | Read_v2 | Agoraphobia without mention of panic attacks |
| E2023 | Read_v3 | Social phobia, fear of eating in public |
| E2023 | Read_v2 | Social phobia, fear of eating in public |
| E2024 | Read_v3 | Social phobia, fear of public speaking |
| E2024 | Read_v2 | Social phobia, fear of public speaking |
| E2025 | Read_v3 | Social phobia, fear of public washing |
| E2025 | Read_v2 | Social phobia, fear of public washing |
| E2026 | Read_v3 | Acrophobia |
| E2026 | Read_v2 | Acrophobia |
| E2027 | Read_v3 | Animal phobia |
| E2027 | Read_v2 | Animal phobia |
| E2028 | Read_v3 | Claustrophobia |
| E2028 | Read_v2 | Claustrophobia |
| E2029 | Read_v3 | Fear of crowds |
| E2029 | Read_v2 | Fear of crowds |
| E202A | Read_v3 | Fear of flying |
| E202A | Read_v2 | Fear of flying |
| E202B | Read_v3 | Cancer phobia |
| E202B | Read_v2 | Cancer phobia |
| E202C | Read_v3 | Dental phobia |
| E202C | Read_v2 | Dental phobia |
| E202D | Read_v3 | Fear of death |
| E202D | Read_v2 | Fear of death |
| E202E | Read_v3 | Fear of pregnancy |
| E202E | Read_v2 | Fear of pregnancy |
| E202z | Read_v3 | (Weight fixation) or (phobic disorder NOS) |
| E202z | Read_v2 | Phobic disorder NOS |
| E28z. | Read_v3 | (Examination fear) or (flying phobia) or (stage fright) or (acute stress reaction NOS) |
| Eu40. | Read_v3 | [X]Phobic anxiety disorders |
| Eu40. | Read_v2 | [X]Phobic anxiety disorders |
| Eu400 | Read_v3 | [X] Agoraphobia (& [without history of panic disorder] or [with panic disorder]) |
| Eu400 | Read_v2 | [X]Agoraphobia |
| Eu401 | Read_v3 | Social phobia |
| Eu401 | Read_v2 | [X]Social phobias |
| Eu402 | Read_v3 | [X] Specific (isolated) phobias (& [acrophobia] or [animal] or [claustrophobia] or [simple phobia]) |
| Eu402 | Read_v2 | [X]Specific (isolated) phobias |
| Eu403 | Read_v3 | Needle phobia |
| Eu403 | Read_v2 | [X]Needle phobia |
| Eu40y | Read_v2 | [X]Other phobic anxiety disorders |
| Eu40z | Read_v3 | [X]Phobic anxiety disorder, unspecified |
| Eu40z | Read_v2 | [X]Phobic anxiety disorder, unspecified |
| Eu41. | Read_v3 | [X]Other anxiety disorders |
| Eu41. | Read_v2 | [X]Other anxiety disorders |
| Eu410 | Read_v3 | [X]Panic disorder [episodic paroxysmal anxiety] |
| Eu410 | Read_v2 | [X]Panic disorder [episodic paroxysmal anxiety] |

| | | |
|---|---|---|
| Eu411 | Read_v3 | Anxiety neurosis |
| Eu411 | Read_v2 | [X]Generalized anxiety disorder |
| Eu413 | Read_v3 | [X]Other mixed anxiety disorders |
| Eu413 | Read_v2 | [X]Other mixed anxiety disorders |
| Eu41y | Read_v3 | [X] Anxiety disorders: [other specified] or [anxiety hysteria] |
| Eu41y | Read_v2 | [X]Other specified anxiety disorders |
| Eu41z | Read_v3 | [X]Anxiety disorder, unspecified |
| Eu41z | Read_v2 | [X]Anxiety disorder, unspecified |
| Ua1qa | Read_v3 | Fear of dentist |
| Ua1qc | Read_v3 | Fear of not coping with treatment |
| Ua1qd | Read_v3 | Fear of lifts |
| Ua1qe | Read_v3 | Fear of thunderstorm |
| Ua1qS | Read_v3 | Panic attack |
| Ua1qs | Read_v3 | Fear of wetting self in public |
| Ua1qt | Read_v3 | Fear of losing control of bowels in public |
| Ua1qU | Read_v3 | Fear of walking |
| Ua1qV | Read_v3 | Fear of mobilising |
| Ua1qW | Read_v3 | Fear of disconnection from ventilator |
| Ua1qX | Read_v3 | Fear of being left alone during period of dependence |
| Ua1qY | Read_v3 | Fear of being left alone |
| X00Sr | Read_v3 | Erythrophobia |
| X00SV | Read_v3 | Agoraphobia |
| X00SW | Read_v3 | Social phobia |
| X00SX | Read_v3 | Specific phobia |
| X00SY | Read_v3 | Needle phobia |
| X00Sy | Read_v3 | Weight fixation |
| X50G2 | Read_v3 | Trichophobia |
| X50G3 | Read_v3 | Parasitophobia |
| X50G5 | Read_v3 | Venereophobia |
| X50G6 | Read_v3 | Syphilophobia |
| X50GI | Read_v3 | Bromisodrophobia |
| X761a | Read_v3 | Fear of swallowing |
| X761b | Read_v3 | Fear of collapsing |
| X761c | Read_v3 | Fear of fainting |
| X761d | Read_v3 | Fear of having a heart attack |
| X761e | Read_v3 | Fear of shaking |
| X761f | Read_v3 | Fear of sweating |
| X761G | Read_v3 | Loss of hope for the future |
| X761g | Read_v3 | Fear of dying |
| X761h | Read_v3 | Fear of going crazy |
| X761i | Read_v3 | Fear of losing emotional control |
| X761j | Read_v3 | Fear of becoming fat |
| X761k | Read_v3 | Anxiety about behaviour or performance |
| X761l | Read_v3 | Fear of appearing ridiculous |
| X761m | Read_v3 | Fear of saying the wrong thing |
| X761n | Read_v3 | Fear of going out |
| X761p | Read_v3 | Fear of empty streets |
| X761q | Read_v3 | Fear of open spaces |
| X761r | Read_v3 | Fear of crossing streets |
| X761R | Read_v3 | Situation avoidance behaviour |
| X761t | Read_v3 | Fear of transport |
| X761T | Read_v3 | Anxiety about body function or health |
| X761U | Read_v3 | Fear of losing control of bowels |
| X761u | Read_v3 | Social fear |
| X761v | Read_v3 | Fear of activities in public |
| X761V | Read_v3 | Fear of wetting self |
| X761w | Read_v3 | Fear of eating in public |
| X761W | Read_v3 | Fear of vomiting in public |
| X761x | Read_v3 | Fear of public speaking |
| X761X | Read_v3 | Fear of having a fit |
| X761y | Read_v3 | Fear of using public toilets |
| X761Y | Read_v3 | Fear of choking |
| X761z | Read_v3 | Fear of writing in public |
| X761Z | Read_v3 | Anxiety about blushing |
| X7620 | Read_v3 | Fear of social group activities |
| X7621 | Read_v3 | Fear of being in a small group |
| X7622 | Read_v3 | Fear of social gatherings |
| X7623 | Read_v3 | Fear of speaking on the phone |
| X7624 | Read_v3 | Fear of speaking to people in authority |
| X7625 | Read_v3 | Fear of being laughed at |
| X7626 | Read_v3 | Fear of being watched |
| X7627 | Read_v3 | Specific fear |
| X7628 | Read_v3 | Fear of natural phenomena |
| X7629 | Read_v3 | Fear of the dark |
| X762a | Read_v3 | Fear of ghosts |

| | | |
|---|---|---|
| X762A | Read_v3 | Fear of animals |
| X762b | Read_v3 | Fear of school |
| X762B | Read_v3 | Fear of feathers |
| X762C | Read_v3 | Fear of enclosed spaces |
| X762E | Read_v3 | Fear of tunnels |
| X762F | Read_v3 | Fear of phone boxes |
| X762G | Read_v3 | Fear of flying |
| X762H | Read_v3 | Flying phobia |
| X762I | Read_v3 | Fear associated with illness and body function |
| X762J | Read_v3 | Fear of anaesthetic |
| X762K | Read_v3 | Fear of general anaesthetic |
| X762L | Read_v3 | Fear of awareness under general anaesthetic |
| X762M | Read_v3 | Fear of not waking from general anaesthetic |
| X762N | Read_v3 | Fear of local anaesthetic |
| X762O | Read_v3 | Fear of problem after anaesthetic |
| X762P | Read_v3 | Fear of needles |
| X762R | Read_v3 | Fear of surgical masks |
| X762S | Read_v3 | Fear of hospitals |
| X762T | Read_v3 | Fear of death |
| X762U | Read_v3 | Fear of contracting disease |
| X762V | Read_v3 | Fear of infection |
| X762W | Read_v3 | Fear of contracting venereal disease |
| X762X | Read_v3 | Fear of contracting HIV infection |
| X762Y | Read_v3 | Fear of contracting radiation sickness |
| X762Z | Read_v3 | Fear of the bogey man |
| Xa00r | Read_v3 | Fear of heights |
| Xa00s | Read_v3 | Fear of water |
| Xa1a8 | Read_v3 | Examination phobia |
| Xa1Ev | Read_v3 | Fear of needles |
| Xa3Vk | Read_v3 | Fear of insects |
| Xa3Vl | Read_v3 | Fear of birds |
| Xa3WI | Read_v3 | Fear of blood |
| Xa3WJ | Read_v3 | Fear of getting cancer |
| Xa7lj | Read_v3 | Cancer phobia |
| XaB96 | Read_v3 | Other phobias |
| XaEKL | Read_v3 | Phonophobia |
| XaIvf | Read_v3 | Fear of falling |
| XaK2c | Read_v3 | H/O: agoraphobia |
| XE1aW | Read_v3 | (Anxiety state (& [states] or [panic attack])) or (pseudocyesis) |
| XE1Y7 | Read_v3 | Panic disorder |
| XE1YA | Read_v3 | Phobic anxiety disorder |
| XE1YB | Read_v3 | Phobic disorder NOS |
| XE1Zj | Read_v3 | [X]Other specified anxiety |

*Note:* Clinical codes from Fabbri et al. (2021), doi: 10.1038/s41380-021-01062-9.

### Supplement 5. Psychotropic medication codes

**Supplement table 3.** Psychotropic medication codes

| UK Biobank code | Drug name |
|---|---|
| 1140879616 | Amitriptyline |
| 1140921600 | Citalopram |
| 1140879540 | Fluoxetine |
| 1140867878 | Sertraline |
| 1140916282 | Venlafaxine |
| 1140909806 | Dosulepin |
| 1140867888 | Paroxetine |
| 1141152732 | Mirtazapine |
| 1141180212 | Escitalopram |
| 1140879634 | Trazodone |
| 1140867876 | Prozac |
| 1140882236 | Seroxat |
| 1141190158 | Cipralex |
| 1141200564 | Duloxetine |
| 1140867726 | Lofepramine |
| 1140879620 | Clomipramine |
| 1140867818 | Nortriptyline |
| 1140879630 | Imipramine |
| 1140879628 | Dothiepin |
| 1141151946 | Cipramil |
| 1140867948 | Amitriptyline |
| 1140867624 | Prothiaden |
| 1140867756 | Trimipramine |
| 1140867884 | Lustral |
| 1141151978 | Reboxetine |
| 1141152736 | Zispin |
| 1141201834 | Cymbalta |
| 1140867690 | Anafranil |
| 1140867640 | Doxepin |
| 1140867920 | Moclobemide |
| 1140867850 | Phenelzine |
| 1140879544 | Fluvoxamine |
| 1141200570 | Yentreve |
| 1140867934 | Triptafen |
| 1140867758 | Surmontil |
| 1140867914 | Tranylcypromine |
| 1140867820 | Allegron |
| 1141151982 | Edronax |
| 1140882244 | Molipaxin |
| 1140879556 | Mianserin |
| 1140867852 | Nardil |
| 1140867860 | Faverin |
| 1140917460 | Nefazodone |
| 1140867938 | Amitriptyline+Chlordiazepoxide |
| 1140867856 | Isocarboxazid |
| 1140867922 | Manerix |
| 1140910820 | Maoi |
| 1140882312 | Sinequan |
| 1140867944 | Tranylcypromine+Trifluoperazine |
| 1140867784 | Ludiomil |
| 1140867812 | Norval |
| 1140867668 | Tryptizol |
| 1140867940* | Fluphenazine hydrochloride+Nortriptyline 1.5mg/30mg tablet |
| 1140867942* | Fluphenazine hcl+Nortriptyline 500micrograms/10mg tablet |
| 1140928916 | Olanzapine |
| 1141152848 | Quetiapine |
| 1140867444 | Risperidone |
| 1140879658 | Chlorpromazine |
| 1140868120 | Trifluoperazine |
| 1141153490 | Amisulpride |
| 1140867304 | Sulpiride |
| 1141152860 | Seroquel |
| 1140867168 | Haloperidol |
| 1141195974 | Aripiprazole |
| 1140867244 | Stelazine |
| 1140867152 | Depixol |
| 1140909800 | Flupentixol |
| 1140867420 | Clozapine |
| 1140879746 | Promazine |
| 1141177762 | Risperdal |

| | |
|---|---|
| 1140867456 | Modecate |
| 1140867952 | Fluanxol |
| 1140867150 | Flupenthixol |
| 1141167976 | Zyprexa |
| 1140882100 | Zuclopenthixol |
| 1140867342 | Clopixol |
| 1140863416 | Largactil |
| 1141202024 | Abilify |
| 1140882098 | Fluphenazine |
| 1140867184 | Haldol |
| 1140867092 | Serenace |
| 1140882320 | Clozaril |
| 1140910358 | Chlorpromazine |
| 1140867208 | Perphenazine |
| 1140909802 | Levomepromazine |
| 1140867134 | Pericyazine |
| 1140867306 | Dolmatil |
| 1140867210 | Fentazin |
| 1140867398 | Fluphenazine |
| 1140867078 | Benperidol |
| 1140867218 | Pimozide |
| 1141201792 | Zaponex |
| 1141200458 | Denzapine |
| 1140867136 | Neulactil |
| 1140879750 | Thioridazine |
| 1140867180 | Dozic |
| 1140867546 | Fluspirilene |
| 1140928260 | Panadeine |
| 1140927956 | Sertindole |
| 1140867490 | Lithium product |
| 1140867494 | Camcolit 250 tablet |
| 1140867498 | Liskonum 450mg m/r tablet |
| 1140867500 | Phasal 300mg m/r tablet |
| 1140867504 | Priadel 200mg m/r tablet |
| 1140867518 | Litarex 564mg m/r tablet |
| 1140867520 | Li-liquid 5.4mmol/5ml oral solution |

*Note:* Adapted from Davis et al. (2019), doi: 10.1002/mpr.1796. *medication code not included in Davis et al. (2019).

**Supplement 6. Smith et al. mood disorder criteria**

**Mood disorder** (adapted from Smith et al. (2013), doi: 10.1371/journal.pone.0075362).

**1. Probable bipolar disorder (type I):**
4642 ever manic/hyper 2 days OR 4653 ever irritable/argumentative for 2 days;
plus at least 3 from 6156.01 (more active), 6156.02 (more talkative), 6156.03 (needed less sleep), and 6156.04 (more creative/more ideas);
plus 5663 duration of a week or more;
plus 5674 needed treatment or caused problems at work.

**2. Probable bipolar disorder (type II):**
4642 ever manic/hyper 2 days OR 4653 ever irritable/argumentative for 2 days;
plus at least 3 from 6156.01 (more active), 6156.02 (more talkative), 6156.03 (needed less sleep), and 6156.04 (more creative/more ideas);
plus 5663 duration of a week or more.

**3. Single probable episode of major depression:**
4598 ever depressed/down for a whole week; plus 4609 at least two weeks duration; plus
4620 only one episode, plus 2090 ever seen a GP or 2100 a psychiatrist for nerves, anxiety, depression
OR
4631 ever anhedonic (unenthusiasm/uninterest) for a whole week; plus 5375 at least two weeks; plus
5386 only one episode; plus 2090 ever seen a GP or 2100 a psychiatrist for nerves, anxiety, depression.

**4. Probable recurrent major depression (moderate):**
4598 ever depressed/down for a whole week; plus 4609 at least two weeks duration; plus 4620 at least two episodes; plus 2090 ever seen a GP (but not a psychiatrist) for nerves, anxiety, depression
OR
4631 ever anhedonic (unenthusiasm/uninterest) for a whole week; plus 5375 at least two weeks; plus
5386 at least two episodes; plus 2090 ever seen a GP (but not a psychiatrist) for nerves, anxiety, depression.

**5. Probable recurrent major depression (severe):**
4598 ever depressed/down for a whole week; plus 4609 at least two weeks duration; plus 4620 at least two episodes; plus 2100 ever seen a psychiatrist for nerves, anxiety, depression
OR
4631 ever anhedonic (unenthusiasm/uninterest) for a whole week; plus 5375 at least two weeks; plus
5386 at least two episodes; plus 2100 ever seen a psychiatrist for nerves, anxiety, depression.

**Supplement 7. Physiological measures**

Description of physiological measures reproduced from Mutz et al. (2021).[1]

**Physiological measures**
All physiological measures were collected by certified healthcare technicians or nurses using a direct data entry system. Participants were asked to remove any outer garments, shoes, socks or tights. The assessment lasted about 10-15 minutes.

*Hand-grip strength*
Hand-grip strength in whole kilogram force units was measured using a Jamar J00105 hydraulic hand dynamometer (measurement range 0-90 kg). Participants were asked to sit upright in a chair and place their forearms on armrests. The dynamometer handle was set to the second incremental slot or, in participants with very large hands, moved to the third slot. Participants kept their elbow adjacent to their side and bent to a 90° angle with their thumb facing upward. A maximal score was obtained from each participant's right and left hand. We used the maximal grip strength of the participant's self-reported dominant hand. If no data on handedness were available, we used the highest value of both hands. This variable has been used previously in UK Biobank research,[2] although other studies have used the highest value of left and right hand[3] or calculated the average grip strength from both hands.[4] We used absolute units because these are the simplest to use in risk assessment. A previous UK Biobank study found no evidence of differences in mortality or disease incidence prediction when grip strength was expressed in absolute terms compared to when expressed relative to anthropometric traits (height, weight, fat-free mass and BMI).[4]

*Body composition*
Weight measurements were obtained with a Tanita BC-418 MA body composition analyser, or, in individuals with pacemaker or females who reported that they were or might be pregnant, using a manual scale. Standing height was measured using a Seca 202 height measure. Body mass index (BMI) was calculated as weight divided by standing height squared ($kg/m^2$). Waist and hip circumference in cm were measured using a Wessex non-stretchable sprung tape. Waist-to-hip ratio was calculated by dividing waist circumference by hip circumference. Whole body fat mass and fat-free mass in kg and body fat percentage (measurement range 1-75%) were estimated by electrical bio-impedance.

*Pulmonary function*
Volumetric measures of lung function were quantified using breath spirometry with a Vitalograph Pneumotrac 6800 following standard procedures. Participants were asked to record two to three blows, each lasting for at least six seconds, within a period of approximately six minutes. The reproducibility of the first two blows was automatically compared and, if acceptable (defined as a ≤5% difference in forced vital capacity (FVC) and forced expiratory volume in one second ($FEV_1$)), a third blow was not required. FVC in litres describes the maximum amount of air that can be exhaled when blowing out as fast as possible after a deep breath. $FEV_1$ in litres describes the amount of air that can be exhaled in one second when blowing out as fast as possible. We used the derived best measure for both FVC and $FEV_1$ which was the maximum value from reproducible spirograms, in line with previous research.[5] Given that these measures are affected by several factors unrelated to pulmonary function (e.g., effort and body size), we also calculated the ratio of $FEV_1$ to FVC. Peak expiratory flow (PEF) in litres per minute represents a person's maximum speed of expiration. PEF is determined by physiological factors such as lung volume and elasticity or expiratory muscle strength. It is used for monitoring asthma and diagnosing chronic obstructive pulmonary disease. We calculated the average of all available readings. Spirometry was not performed in participants who confirmed or were unsure that they had any of the following contra-indications: chest infection in the last month (i.e., influenza, bronchitis, severe cold and pneumonia); history of a detached retina; heart attack or surgery to eyes, chest or abdomen in the last three months; history of a collapsed lung; pregnancy in the 1st or 3rd trimester; currently on medication for tuberculosis.

*Heel bone mineral density*
Heel bone mineral density was estimated by quantitative ultrasound assessment of the calcaneus using a Sahara Clinical Bone Sonometer. Participants were asked to sit with their back straight and had their left heel measured first. Measurement of both heels was performed only during later stages of the baseline assessment. Measures of speed of sound in metres per second and broadband ultrasound attenuation (decibels/megahertz) were combined into a quantitative ultrasound index. From this an estimate of heel bone mineral density in grams per $cm^2$ was derived based on the assumption that sound waves travel differently through denser bones.

**Cardiovascular measures**
Blood pressure, pulse rate and arterial stiffness (pulse wave velocity) were measured by trained nurses during a separate stage of the baseline assessment.

*Blood pressure*
Seated systolic and diastolic resting blood pressure in millimetres of mercury (mmHg) was measured twice using an Omron 705 IT digital blood pressure monitor (measurement range 0-255 mmHg) following standard procedures. Participants were asked to loosen or remove any restrictive clothing and put their arm on a desktop. Measurements were taken from the left upper arm or, if impractical, from the right arm. If there was a problem with the measurement, it was repeated. Three different cuff sizes were available and a Seca tape was used to determine the circumference of the midpoint of the upper arm. If the largest cuff size was too small or the digital blood pressure monitor did not produce a reading, a manual sphygmomanometer was used in conjunction with a stethoscope. A second measurement was taken at least one minute after the first measure, following the same procedure. We used an average of the two readings to reduce potential measurement error.

*Pulse rate*
Resting pulse rate in beats per minute was recorded during the blood pressure measurements using the Omron 705 IT device or, exceptionally, a manual sphygmomanometer. We used an average of the two readings to reduce potential measurement error.

*Arterial stiffness*
Arterial stiffness (vascular reactivity) is an independent predictor of cardiovascular risk and mortality that can be measured rapidly, inexpensively, and without special training.[6] Resting pulse wave velocity was measured non-invasively using finger photoplethysmography with a PulseTrace PCA2 infra-red sensor. The pulse waveform was recorded over a period of 10-15 seconds, with the sensor clipped to the end of the index finger of the non-dominant hand while the participant was sitting. If the waveform did not fill at least 2/3 of the display of the device, or did not stabilize within one minute, the measurement was repeated on a larger finger or on the thumb. The shape of the waveform is directly related to the time it takes for the pulse wave to travel through the arterial tree in the lower body and to be reflected back to the finger. The time between the peaks of the waveform (the pulse wave peak-to-peak time, i.e., the difference between the peak values of direct and reflected components) was divided by the participant's height to obtain the arterial stiffness index in metres per second. A higher score on the index represents stiffer arteries. The method has been externally validated and is highly correlated with the gold-standard carotid-femoral pulse wave velocity.[7]

### Supplement 8. Additional results

#### Study population

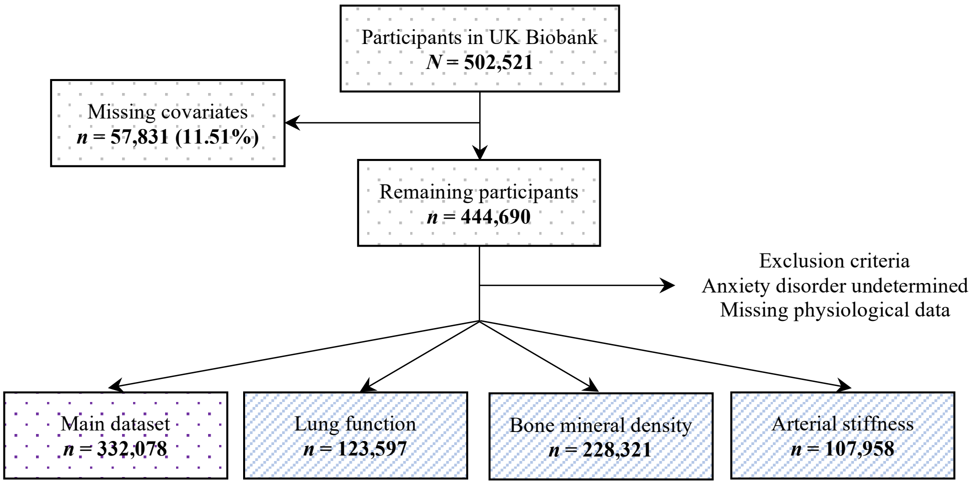

**Supplement figure 1.** Study population. Physiological measures included in the main dataset were hand-grip strength, blood pressure, pulse rate and measures of body composition.

### Sample characteristics

**Supplement table 4.** Sample characteristics

| | | Female | | Male | |
|---|---|---|---|---|---|
| | Overall (N=502521) | Healthy control (N=145364) | Anxiety disorder (N=29482) | Healthy control (N=141992) | Anxiety disorder (N=15240) |
| **Age** | | | | | |
| Mean (SD) | 56.53 (8.10) | 56.44 (8.03) | 55.40 (7.78) | 56.55 (8.26) | 56.00 (7.93) |
| **Ethnicity** | | | | | |
| White | 472711 (94.1%) | 137745 (94.8%) | 28667 (97.2%) | 135015 (95.1%) | 14813 (97.2%) |
| Mixed-race | 2958 (0.6%) | 929 (0.6%) | 172 (0.6%) | 648 (0.5%) | 68 (0.4%) |
| Black | 8061 (1.6%) | 2524 (1.7%) | 234 (0.8%) | 2049 (1.4%) | 68 (0.4%) |
| Asian | 9882 (2.0%) | 2249 (1.5%) | 186 (0.6%) | 2834 (2.0%) | 192 (1.3%) |
| Chinese | 1574 (0.3%) | 620 (0.4%) | 34 (0.1%) | 387 (0.3%) | 22 (0.1%) |
| Other | 4558 (0.9%) | 1297 (0.9%) | 189 (0.6%) | 1059 (0.7%) | 77 (0.5%) |
| Prefer not to answer | 1662 (0.3%) | | | | |
| Do not know | 217 (0.0%) | | | | |
| Missing | 898 (0.2%) | | | | |
| **Highest qualification** | | | | | |
| None | 85274 (17.0%) | 21710 (14.9%) | 3437 (11.7%) | 21095 (14.9%) | 1859 (12.2%) |
| O levels/GCSEs/CSEs | 132086 (26.3%) | 43016 (29.6%) | 8394 (28.5%) | 34999 (24.6%) | 3539 (23.2%) |
| A levels/NVQ/HND/HNC[1] | 113859 (22.7%) | 32548 (22.4%) | 6926 (23.5%) | 34655 (24.4%) | 3754 (24.6%) |
| Degree | 161168 (32.1%) | 48090 (33.1%) | 10725 (36.4%) | 51243 (36.1%) | 6088 (39.9%) |
| Prefer not to answer | 5493 (1.1%) | | | | |
| Missing | 4641 (0.9%) | | | | |
| **Walking[2]** | | | | | |
| Mean (SD) | 5.39 (1.93) | 5.50 (1.83) | 5.35 (1.95) | 5.33 (1.98) | 5.22 (2.05) |
| Prefer not to answer | 979 (0.2%) | | | | |
| Unable to walk | 1929 (0.4%) | | | | |
| Do not know | 6687 (1.3%) | | | | |
| Missing | 874 (0.2%) | | | | |
| **Moderate activity[2]** | | | | | |
| Mean (SD) | 3.63 (2.33) | 3.66 (2.31) | 3.54 (2.35) | 3.62 (2.31) | 3.45 (2.34) |
| Prefer not to answer | 2273 (0.5%) | | | | |
| Do not know | 24120 (4.8%) | | | | |
| Missing | 878 (0.2%) | | | | |
| **Vigorous activity[2]** | | | | | |
| Mean (SD) | 1.84 (1.96) | 1.74 (1.86) | 1.61 (1.82) | 2.11 (2.03) | 1.98 (2.01) |
| Prefer not to answer | 4116 (0.8%) | | | | |
| Do not know | 22582 (4.5%) | | | | |
| Missing | 878 (0.2%) | | | | |
| **Smoking status** | | | | | |
| Never | 273528 (54.4%) | 90936 (62.6%) | 16356 (55.5%) | 73322 (51.6%) | 7040 (46.2%) |
| Former | 173064 (34.4%) | 44022 (30.3%) | 10308 (35.0%) | 53227 (37.5%) | 6202 (40.7%) |
| Current | 52979 (10.5%) | 10406 (7.2%) | 2818 (9.6%) | 15443 (10.9%) | 1998 (13.1%) |
| Prefer not to answer | 2059 (0.4%) | | | | |
| Missing | 891 (0.2%) | | | | |
| **Alcohol intake frequency** | | | | | |
| Never | 40645 (8.1%) | 11483 (7.9%) | 2609 (8.8%) | 6971 (4.9%) | 1124 (7.4%) |
| Special occasions | 58011 (11.5%) | 19697 (13.6%) | 4311 (14.6%) | 9157 (6.4%) | 1102 (7.2%) |
| 1-3/month | 55856 (11.1%) | 18469 (12.7%) | 3772 (12.8%) | 12207 (8.6%) | 1404 (9.2%) |
| 1-2/week | 129294 (25.7%) | 38947 (26.8%) | 7293 (24.7%) | 37266 (26.2%) | 3620 (23.8%) |
| 3-4/week | 115443 (23.0%) | 32344 (22.3%) | 6262 (21.2%) | 39309 (27.7%) | 3776 (24.8%) |
| Daily/almost daily | 101770 (20.3%) | 24424 (16.8%) | 5235 (17.8%) | 37082 (26.1%) | 4214 (27.7%) |
| Prefer not to answer | 605 (0.1%) | | | | |
| Missing | 897 (0.2%) | | | | |
| **Sleep duration** | | | | | |
| Mean (SD) | 7.15 (1.11) | 7.18 (1.02) | 7.16 (1.18) | 7.13 (1.00) | 7.11 (1.16) |
| Prefer not to answer | 386 (0.1%) | | | | |
| Do not know | 2943 (0.6%) | | | | |
| Missing | 887 (0.2%) | | | | |
| **Antihypertensive use** | | | | | |
| Yes | 104001 (20.70%) | 23649 (16.3%) | 4749 (16.1%) | 31565 (22.2%) | 3919 (25.7%) |
| No | 389915 (77.59%) | 121715 (83.7%) | 24733 (83.9%) | 110427 (77.8%) | 11321 (74.3%) |
| Missing | 8605 (1.71%) | | | | |

*Note:* Descriptive statistics for covariates based on main dataset *N*=332,078. GCSEs = general certificate of secondary education; CSE = certificate of secondary education; NVQ = national vocational qualification; HND = higher national diploma; HNC = higher national certificate. [1]also includes 'other professional qualifications'. [2]number of days per week engaging in these activities for 10+ minutes continuously.

### Case-control numbers - additional analyses

**Supplement table 5.** Case-control numbers for additional analyses

| | | Chronic and/or severe anxiety disorder | | Anxiety disorder without depression | |
|---|---|---|---|---|---|
| Dataset | Sex | Healthy control | Anxiety disorder | Healthy control | Anxiety disorder |
| Main dataset | Female | 145364 (98.22%) | 2628 (1.78%) | 145364 (93.64%) | 9865 (6.36%) |
| | Male | 141992 (98.96%) | 1491 (1.04%) | 141992 (95.87%) | 6110 (4.13%) |
| Lung function | Female | 50729 (98.36%) | 848 (1.64%) | 50729 (93.71%) | 3407 (6.29%) |
| | Male | 56695 (98.96%) | 597 (1.04%) | 56695 (95.82%) | 2474 (4.18%) |
| Bone mineral density | Female | 101279 (98.39%) | 1655 (1.61%) | 101279 (93.33%) | 7239 (6.67%) |
| | Male | 97749 (98.98%) | 1007 (1.02%) | 97749 (95.60%) | 4503 (4.40%) |
| Arterial stiffness | Female | 45320 (97.83%) | 1006 (2.17%) | 45320 (94.27%) | 2754 (5.73%) |
| | Male | 46549 (98.90%) | 517 (1.10%) | 46549 (96.44%) | 1717 (3.56%) |

*Note:* Percentages correspond to the sex-specific prevalence of cases and controls, separately for the additional analyses of chronic and/or sever anxiety disorder and anxiety disorder without comorbid depression.

### Case-control differences – chronic and/or severe anxiety disorders

**Supplement table 6.** Differences in physiological measures between individuals with chronic and/or severe anxiety disorders and healthy controls

| | Female | | | | | Male | | | | |
|---|---|---|---|---|---|---|---|---|---|---|
| Variable | SMD | 95% CI | | $p_{Bonf.}$ | $p_{BH}$ | SMD | 95% CI | | $p_{Bonf.}$ | $p_{BH}$ |
| Hand-grip strength | -0.075 | -0.114 | -0.037 | 0.005 | 0.001 | -0.075 | -0.127 | -0.024 | 0.088 | 0.017 |
| Systolic blood pressure | -0.198 | -0.237 | -0.160 | <0.001 | <0.001 | -0.153 | -0.204 | -0.102 | <0.001 | <0.001 |
| Diastolic blood pressure | -0.060 | -0.098 | -0.021 | 0.038 | 0.003 | -0.013 | -0.064 | 0.038 | >0.999 | 0.713 |
| Pulse rate | 0.079 | 0.040 | 0.117 | 0.002 | <0.001 | 0.130 | 0.079 | 0.181 | <0.001 | <0.001 |
| Body mass index | 0.133 | 0.094 | 0.172 | <0.001 | <0.001 | 0.078 | 0.027 | 0.129 | 0.100 | 0.017 |
| Body fat percentage | 0.113 | 0.074 | 0.151 | <0.001 | <0.001 | 0.057 | 0.006 | 0.108 | 0.592 | 0.074 |
| Fat mass | 0.158 | 0.120 | 0.197 | <0.001 | <0.001 | 0.083 | 0.032 | 0.134 | 0.057 | 0.014 |
| Fat-free mass | 0.121 | 0.083 | 0.160 | <0.001 | <0.001 | 0.050 | -0.001 | 0.101 | >0.999 | 0.104 |
| Waist-hip ratio | 0.054 | 0.016 | 0.093 | 0.105 | 0.009 | 0.118 | 0.067 | 0.169 | <0.001 | <0.001 |
| Peak expiratory flow | 0.107 | 0.039 | 0.175 | 0.032 | 0.003 | -0.045 | -0.126 | 0.036 | >0.999 | 0.415 |
| Forced expiratory volume 1s | 0.119 | 0.052 | 0.187 | 0.013 | 0.002 | 0.000 | -0.081 | 0.081 | >0.999 | 0.999 |
| Forced vital capacity | 0.109 | 0.041 | 0.177 | 0.035 | 0.003 | 0.018 | -0.063 | 0.099 | >0.999 | 0.713 |
| $FEV_1$ / FVC | 0.055 | -0.013 | 0.123 | >0.999 | 0.163 | -0.044 | -0.124 | 0.037 | >0.999 | 0.437 |
| Heel bone mineral density | 0.029 | -0.019 | 0.078 | >0.999 | 0.234 | -0.075 | -0.137 | -0.013 | 0.324 | 0.046 |
| Arterial stiffness | -0.029 | -0.091 | 0.033 | >0.999 | 0.186 | -0.048 | -0.135 | 0.038 | 0.997 | 0.104 |

*Note:* SMD = standardised mean difference; CI = confidence interval; Bonf. = Bonferroni; BH = Benjamini & Hochberg; $FEV_1$ = forced expiratory volume in one second; FVC = forced vital capacity. *P*-values for Welch's t-test. Negative values correspond to lower values in anxiety disorder cases.

### Case-control differences – anxiety disorders without comorbid depression

**Supplement table 7.** Differences in physiological measures between individuals with anxiety disorders without comorbid depression and healthy controls

| | Female | | | | | Male | | | | |
|---|---|---|---|---|---|---|---|---|---|---|
| Variable | SMD | 95% CI | | $p_{Bonf.}$ | $p_{BH}$ | SMD | 95% CI | | $p_{Bonf.}$ | $p_{BH}$ |
| Hand-grip strength | -0.028 | -0.048 | -0.008 | 0.118 | 0.017 | -0.059 | -0.085 | -0.034 | <0.001 | <0.001 |
| Systolic blood pressure | -0.028 | -0.048 | -0.007 | 0.104 | 0.017 | -0.038 | -0.063 | -0.012 | 0.055 | 0.008 |
| Diastolic blood pressure | -0.032 | -0.053 | -0.012 | 0.027 | 0.007 | -0.029 | -0.055 | -0.004 | 0.354 | 0.044 |
| Pulse rate | 0.039 | 0.018 | 0.059 | 0.005 | 0.002 | 0.049 | 0.023 | 0.075 | 0.004 | 0.001 |
| Body mass index | -0.047 | -0.067 | -0.026 | <0.001 | <0.001 | -0.058 | -0.084 | -0.033 | <0.001 | <0.001 |
| Body fat percentage | -0.019 | -0.040 | 0.001 | >0.999 | 0.101 | 0.007 | -0.018 | 0.033 | >0.999 | 0.706 |
| Fat mass | -0.026 | -0.046 | -0.005 | 0.208 | 0.023 | -0.014 | -0.039 | 0.012 | >0.999 | 0.480 |
| Fat-free mass | -0.049 | -0.069 | -0.028 | <0.001 | <0.001 | -0.066 | -0.092 | -0.041 | <0.001 | <0.001 |
| Waist-hip ratio | 0.005 | -0.016 | 0.025 | >0.999 | 0.675 | 0.040 | 0.015 | 0.066 | 0.032 | 0.005 |
| Peak expiratory flow | -0.016 | -0.050 | 0.019 | >0.999 | 0.489 | -0.020 | -0.060 | 0.020 | >0.999 | 0.480 |
| Forced expiratory volume 1s | -0.014 | -0.049 | 0.020 | >0.999 | 0.491 | -0.010 | -0.051 | 0.030 | >0.999 | 0.706 |
| Forced vital capacity | 0.008 | -0.027 | 0.042 | >0.999 | 0.675 | -0.007 | -0.048 | 0.033 | >0.999 | 0.771 |
| $FEV_1$ / FVC | -0.048 | -0.082 | -0.013 | 0.156 | 0.019 | -0.010 | -0.051 | 0.030 | >0.999 | 0.706 |
| Heel bone mineral density | -0.036 | -0.060 | -0.012 | 0.043 | 0.009 | -0.048 | -0.078 | -0.018 | 0.026 | 0.005 |
| Arterial stiffness | -0.019 | -0.058 | 0.019 | >0.999 | 0.251 | 0.000 | -0.048 | 0.048 | >0.999 | 0.987 |

*Note:* SMD = standardised mean difference; CI = confidence interval; Bonf. = Bonferroni; BH = Benjamini & Hochberg; $FEV_1$ = forced expiratory volume in one second; FVC = forced vital capacity. *P*-values for Welch's t-test. Negative values correspond to lower values in anxiety disorder cases.

**Difference smooths females**

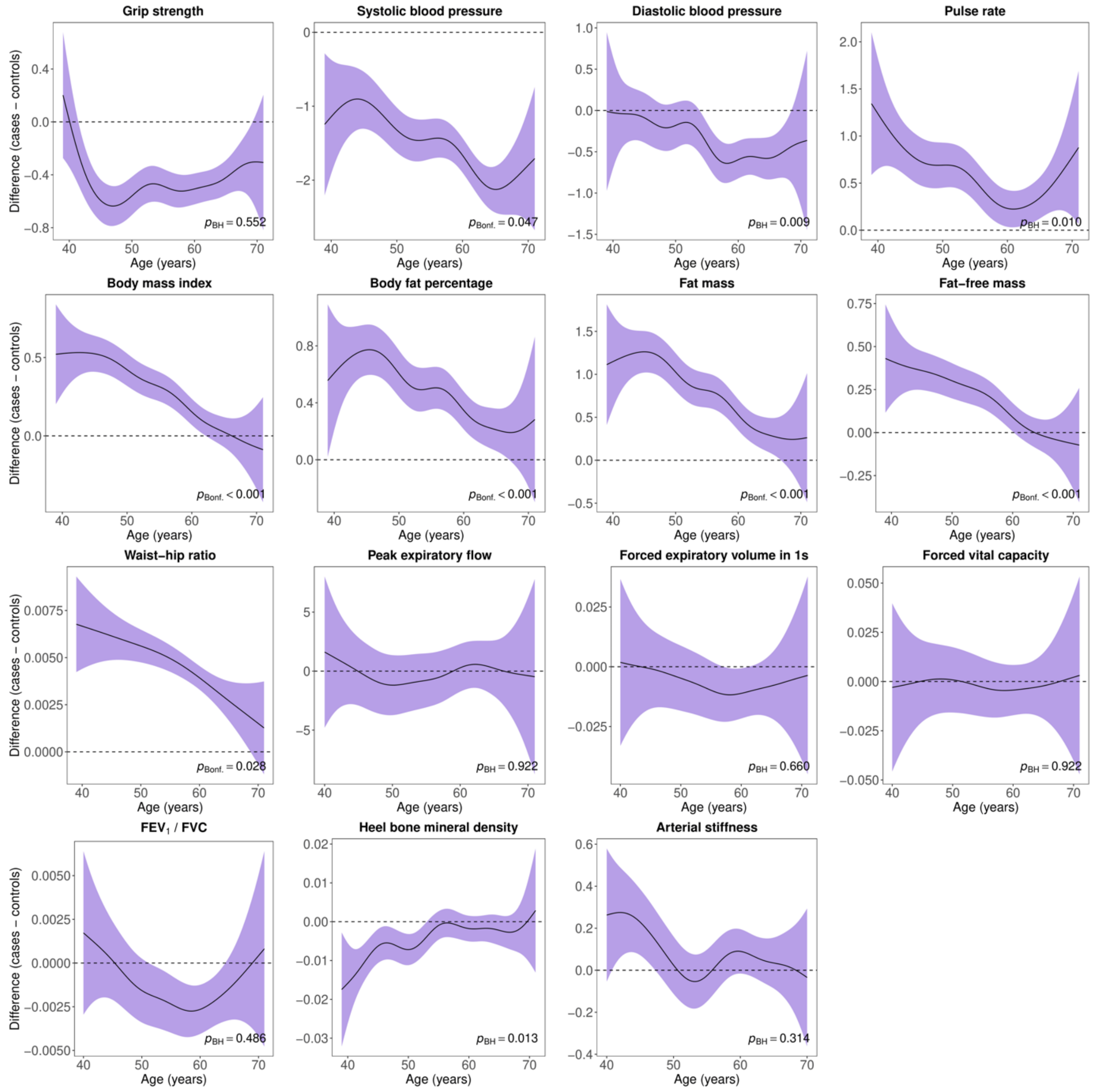

**Supplement figure 2.** Difference smooths comparing age-related changes in physiological measures of females with anxiety disorders to healthy controls. The shaded areas correspond to approximate 95% confidence intervals (± 2 × standard error). Negative values on the y-axes correspond to lower values in females with anxiety disorders compared to healthy controls. The horizontal lines represent no difference between female cases and controls. $FEV_1$ = forced expiratory volume in one second; FVC = forced vital capacity; Bonf. = Bonferroni; BH = Benjamini & Hochberg.

**Difference smooths males**

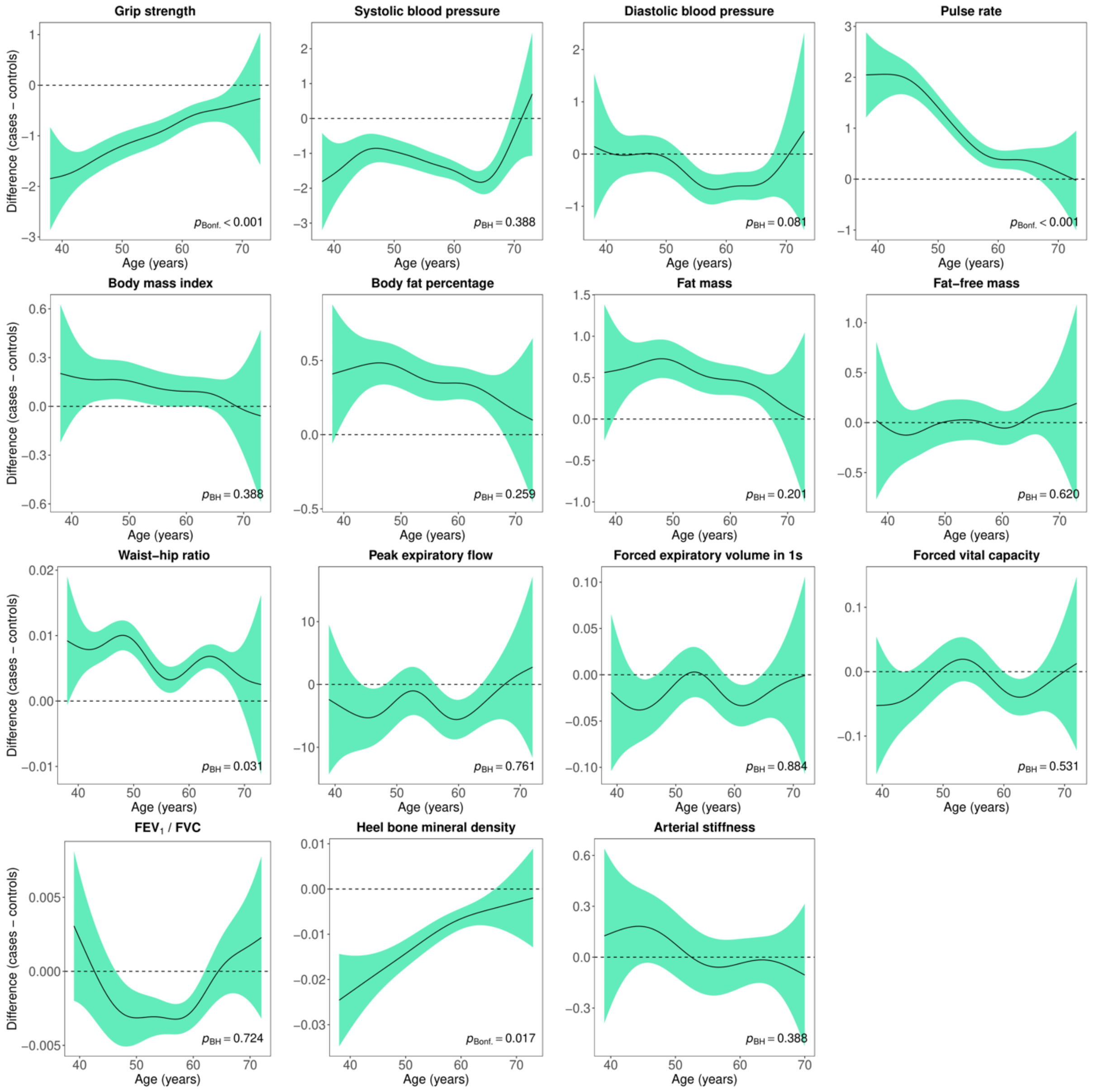

**Supplement figure 3.** Difference smooths comparing age-related changes in physiological measures of males with anxiety disorders to healthy controls. The shaded areas correspond to approximate 95% confidence intervals (± 2 × standard error). Negative values on the y-axes correspond to lower values in males with anxiety disorders compared to healthy controls. The horizontal lines represent no difference between male cases and controls. $FEV_1$ = forced expiratory volume in one second; FVC = forced vital capacity; Bonf. = Bonferroni; BH = Benjamini & Hochberg.

**Adjusted GAMs females**

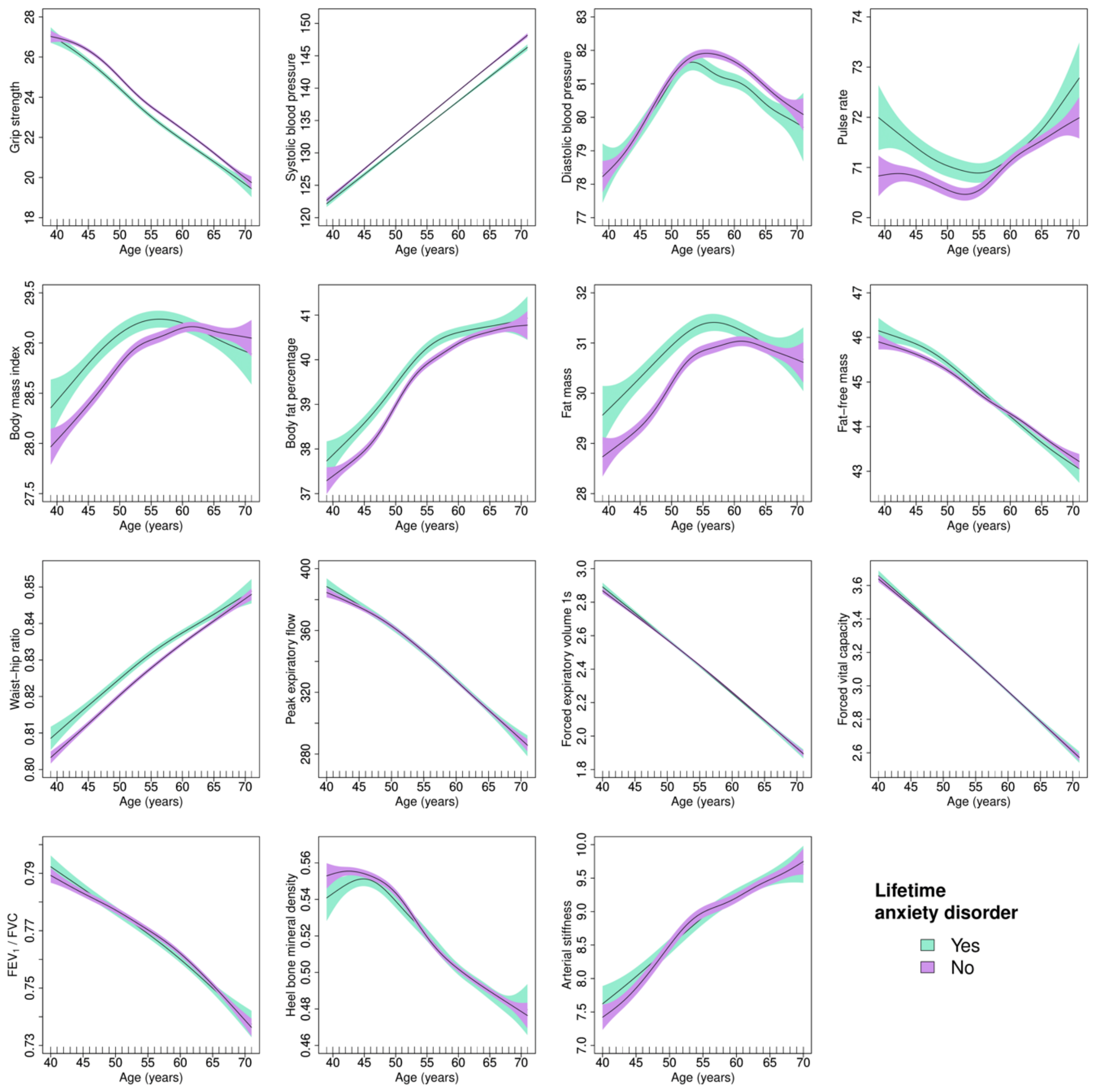

**Supplement figure 4.** Adjusted generalised additive models of age-related changes in physiological measures in females with anxiety disorders and healthy controls. Models were adjusted for ethnicity (except lung function), highest educational/professional qualification, physical activity, smoking status, alcohol intake frequency, sleep duration and, for cardiovascular measures, current use of antihypertensive medications. The solid lines represent physiological measures against smoothing functions of age. The shaded areas correspond to approximate 95% confidence intervals (± 2 × standard error). $FEV_1$ = forced expiratory volume in one second; FVC = forced vital capacity.

**Adjusted difference smooths females**

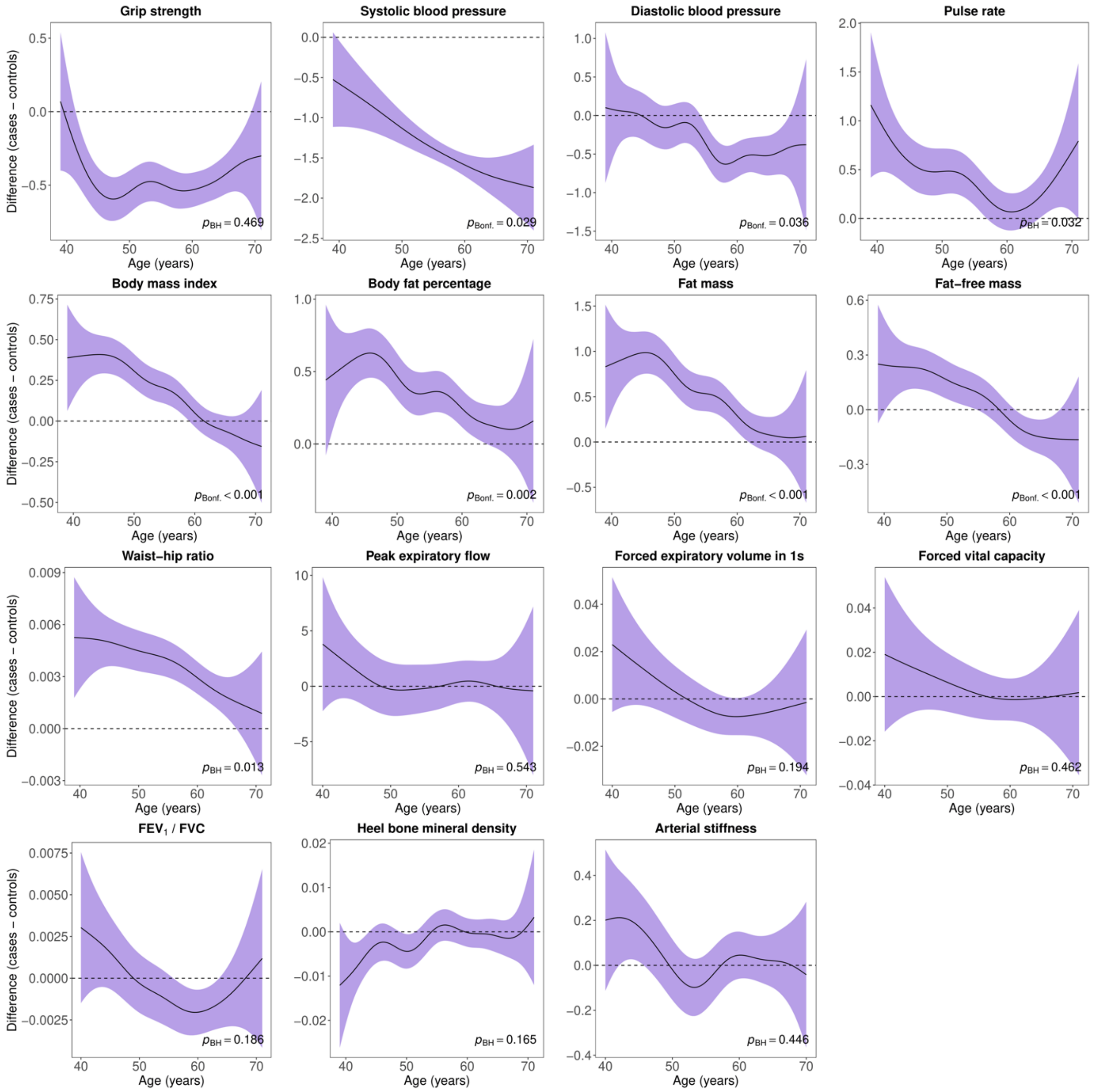

**Supplement figure 5.** Difference smooths comparing age-related changes in physiological measures of females with anxiety disorders to healthy controls. Models were adjusted for ethnicity (except lung function), highest educational/professional qualification, physical activity, smoking status, alcohol intake frequency, sleep duration and, for cardiovascular measures, current use of antihypertensive medications. The shaded areas correspond to approximate 95% confidence intervals (± 2 × standard error). Negative values on the y-axes correspond to lower values in females with anxiety disorders compared to healthy controls. The horizontal lines represent no difference between female cases and controls. $FEV_1$ = forced expiratory volume in one second; FVC = forced vital capacity; Bonf. = Bonferroni; BH = Benjamini & Hochberg.

**Adjusted GAMs males**

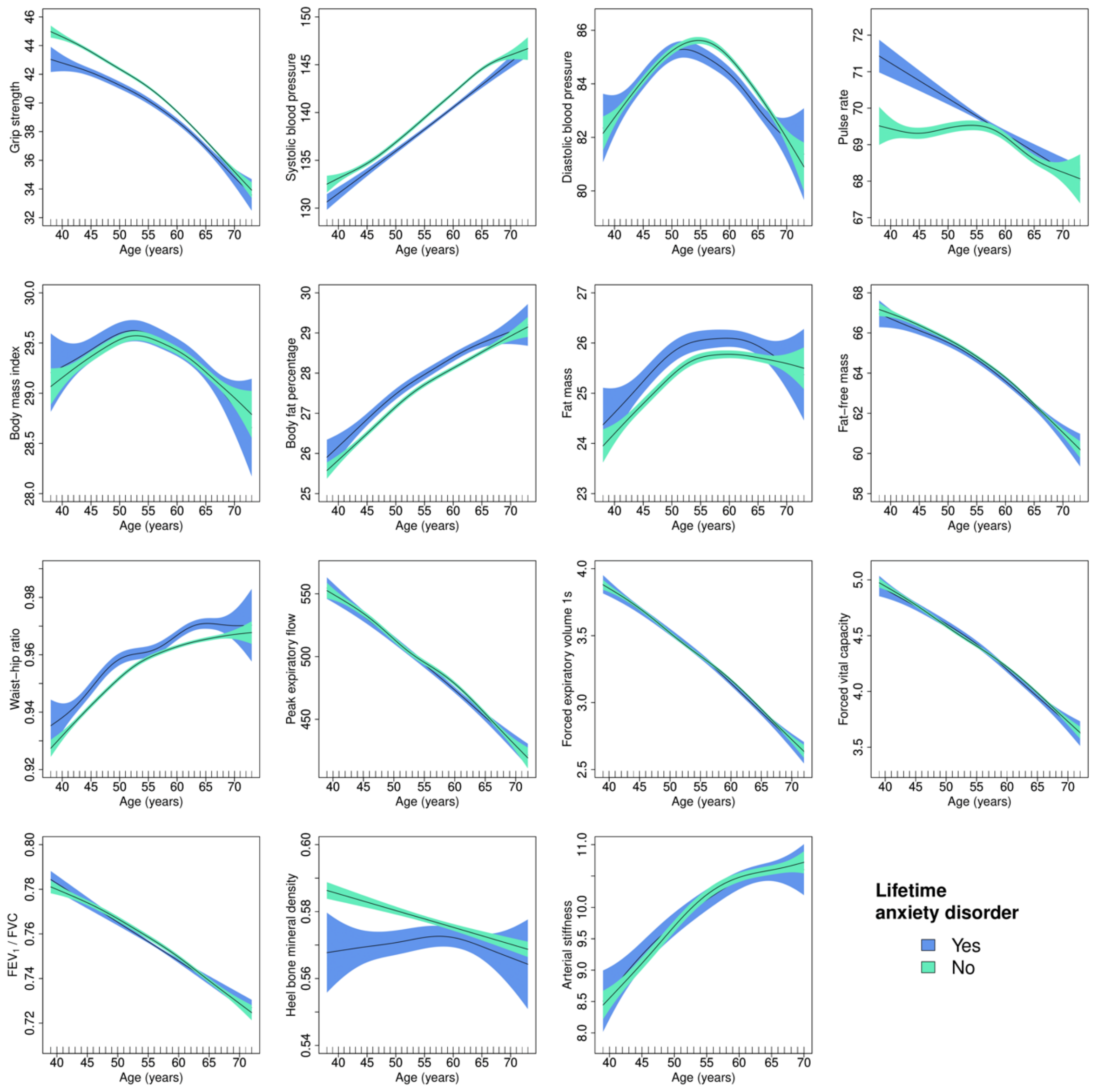

**Supplement figure 6.** Adjusted generalised additive models of age-related changes in physiological measures in males with anxiety disorders and healthy controls. Models were adjusted for ethnicity (except lung function), highest educational/professional qualification, physical activity, smoking status, alcohol intake frequency, sleep duration and, for cardiovascular measures, current use of antihypertensive medications. The solid lines represent physiological measures against smoothing functions of age. The shaded areas correspond to approximate 95% confidence intervals (± 2 × standard error). $FEV_1$ = forced expiratory volume in one second; FVC = forced vital capacity.

**Adjusted difference smooths males**

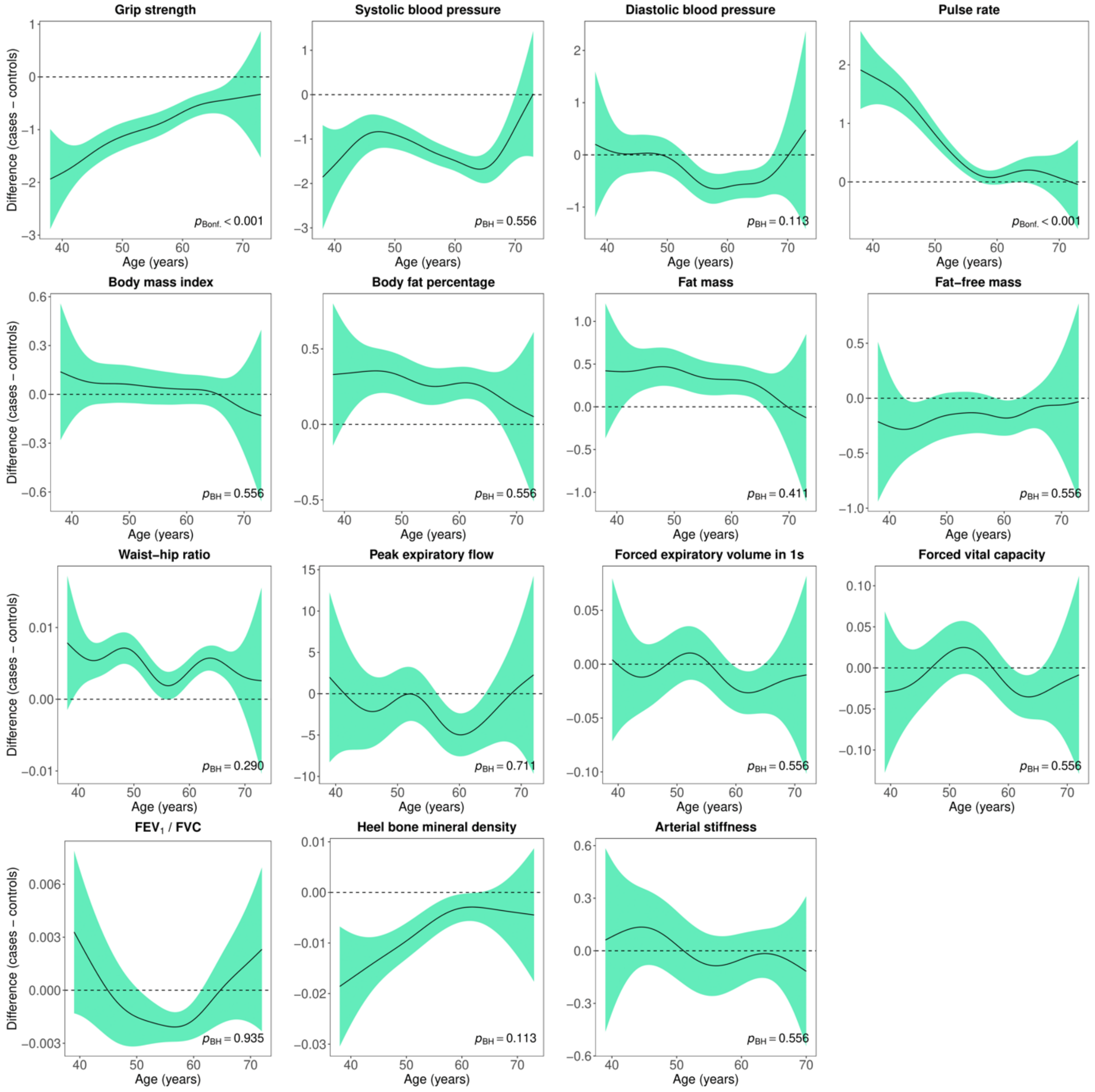

**Supplement figure 7.** Difference smooths comparing age-related changes in physiological measures of males with anxiety disorders to healthy controls. Models were adjusted for ethnicity (except lung function), highest educational/professional qualification, physical activity, smoking status, alcohol intake frequency, sleep duration and, for cardiovascular measures, current use of antihypertensive medications. The shaded areas correspond to approximate 95% confidence intervals (± 2 × standard error). Negative values on the y-axes correspond to lower values in females with anxiety disorders compared to healthy controls. The horizontal lines represent no difference between female cases and controls. $FEV_1$ = forced expiratory volume in one second; FVC = forced vital capacity; Bonf. = Bonferroni; BH = Benjamini & Hochberg.

**Adjusted GAMs females – chronic and/or severe anxiety disorders**

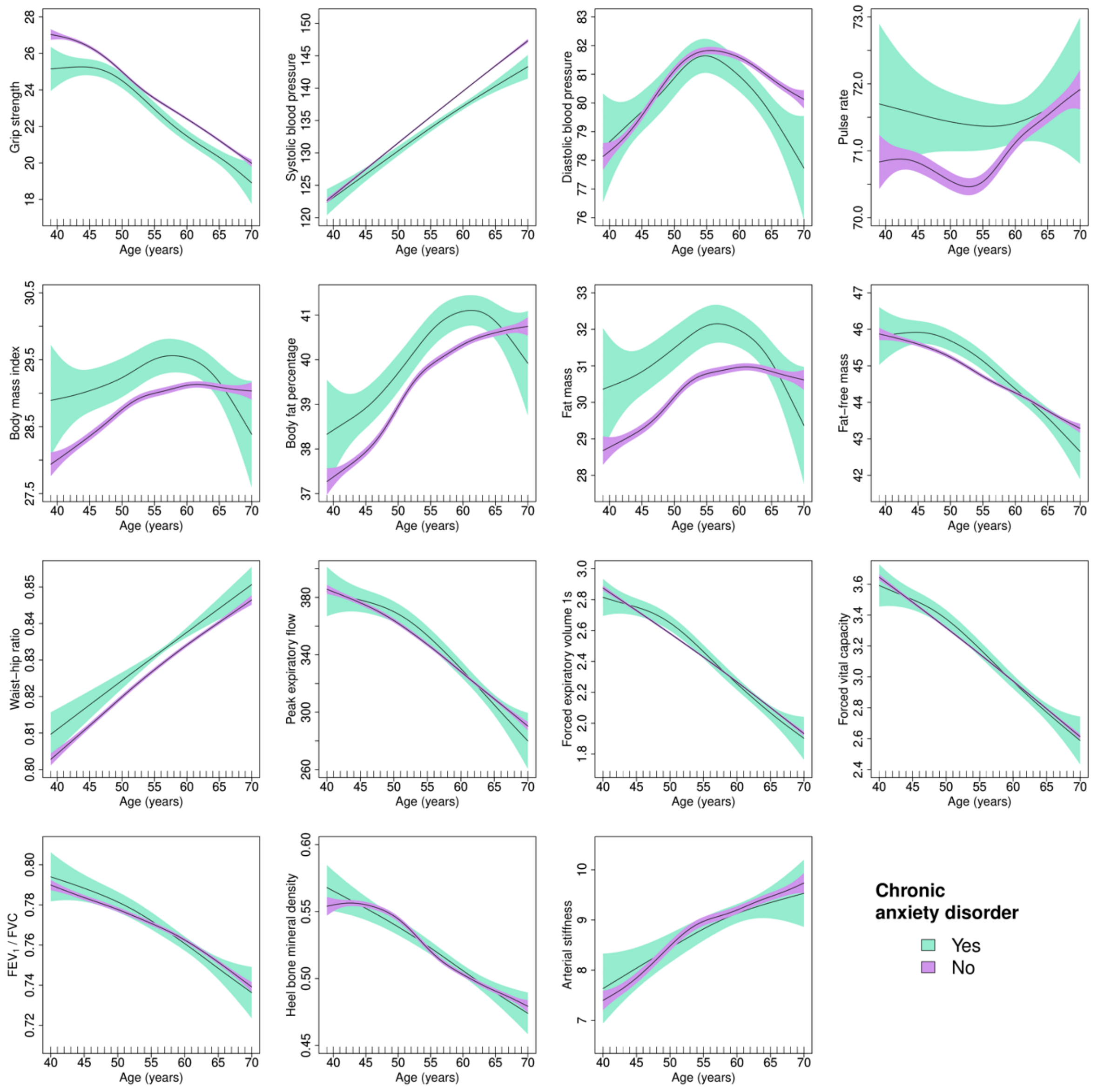

**Supplement figure 8.** Adjusted generalised additive models of age-related changes in physiological measures in females with chronic and/or severe anxiety disorders and healthy controls. Models were adjusted for ethnicity (except lung function), highest educational/professional qualification, physical activity, smoking status, alcohol intake frequency, sleep duration and, for cardiovascular measures, current use of antihypertensive medications. The solid lines represent physiological measures against smoothing functions of age. The shaded areas correspond to approximate 95% confidence intervals (± 2 × standard error). $FEV_1$ = forced expiratory volume in one second; FVC = forced vital capacity.

**Adjusted difference smooths females – chronic and/or severe anxiety disorders**

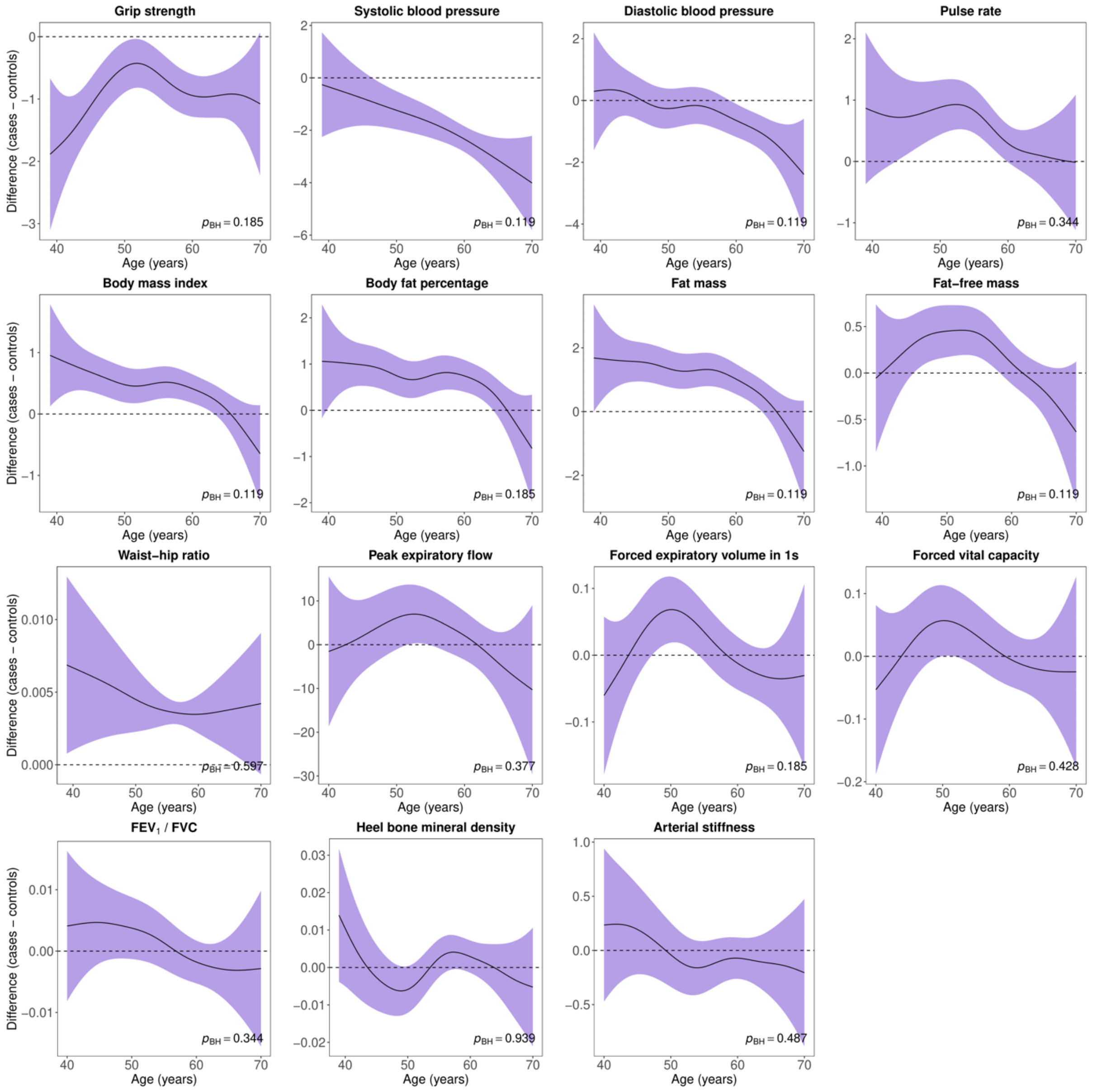

**Supplement figure 9.** Difference smooths comparing age-related changes in physiological measures of females with chronic and/or severe anxiety disorders to healthy controls. Models were adjusted for ethnicity (except lung function), highest educational/professional qualification, physical activity, smoking status, alcohol intake frequency, sleep duration and, for cardiovascular measures, current use of antihypertensive medications. The shaded areas correspond to approximate 95% confidence intervals (± 2 × standard error). Negative values on the y-axes correspond to lower values in females with chronic and/or severe anxiety disorders compared to healthy controls. The horizontal lines represent no difference between female cases and controls. $FEV_1$ = forced expiratory volume in one second; FVC = forced vital capacity; Bonf. = Bonferroni; BH = Benjamini & Hochberg.

**Adjusted GAMs males – chronic and/or severe anxiety disorders**

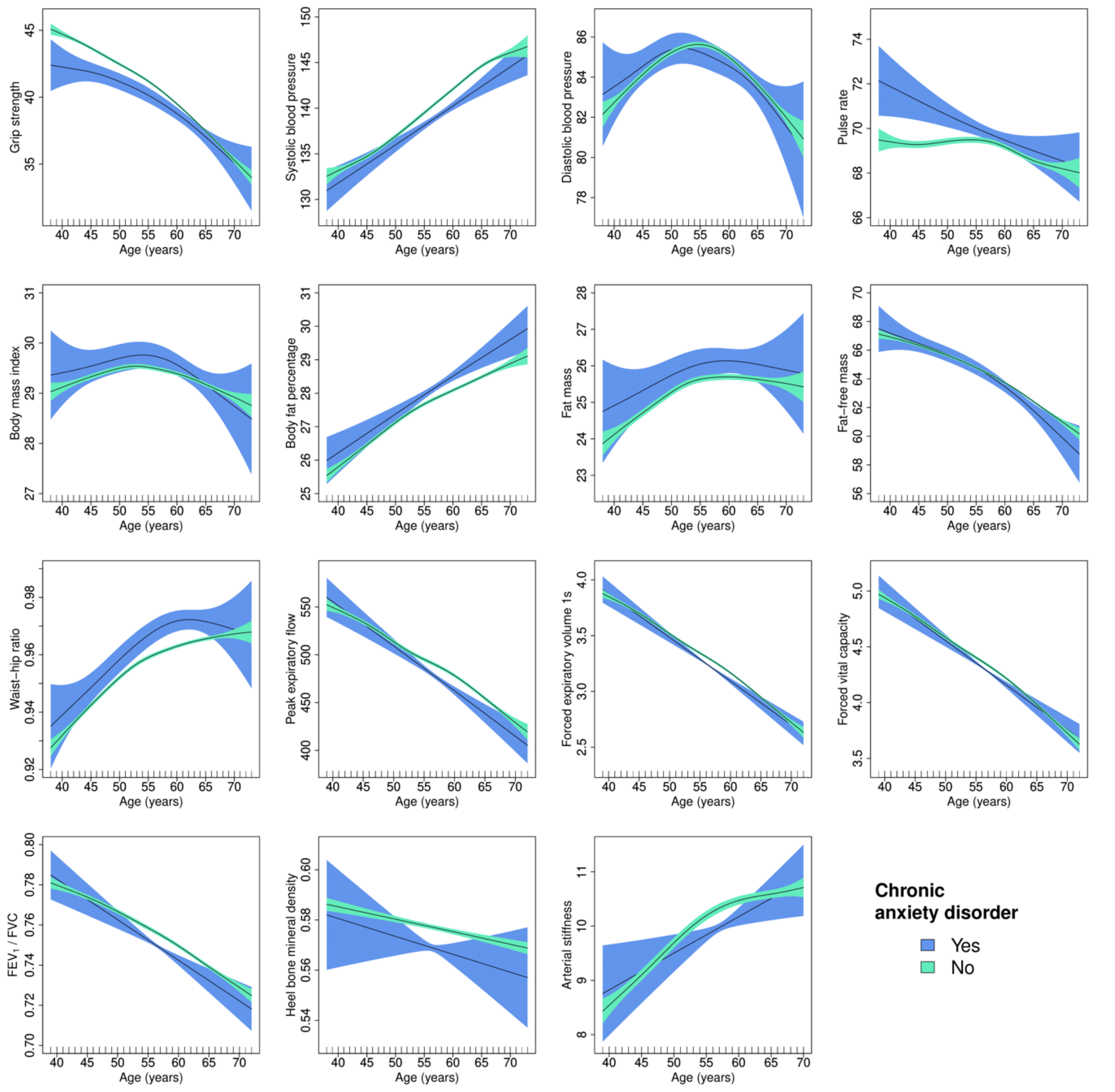

**Supplement figure 10.** Adjusted generalised additive models of age-related changes in physiological measures in males with chronic and/or severe anxiety disorders and healthy controls. Models were adjusted for ethnicity (except lung function), highest educational/professional qualification, physical activity, smoking status, alcohol intake frequency, sleep duration and, for cardiovascular measures, current use of antihypertensive medications. The solid lines represent physiological measures against smoothing functions of age. The shaded areas correspond to approximate 95% confidence intervals (± 2 × standard error). $FEV_1$ = forced expiratory volume in one second; FVC = forced vital capacity.

**Adjusted difference smooths males – chronic and/or severe anxiety disorders**

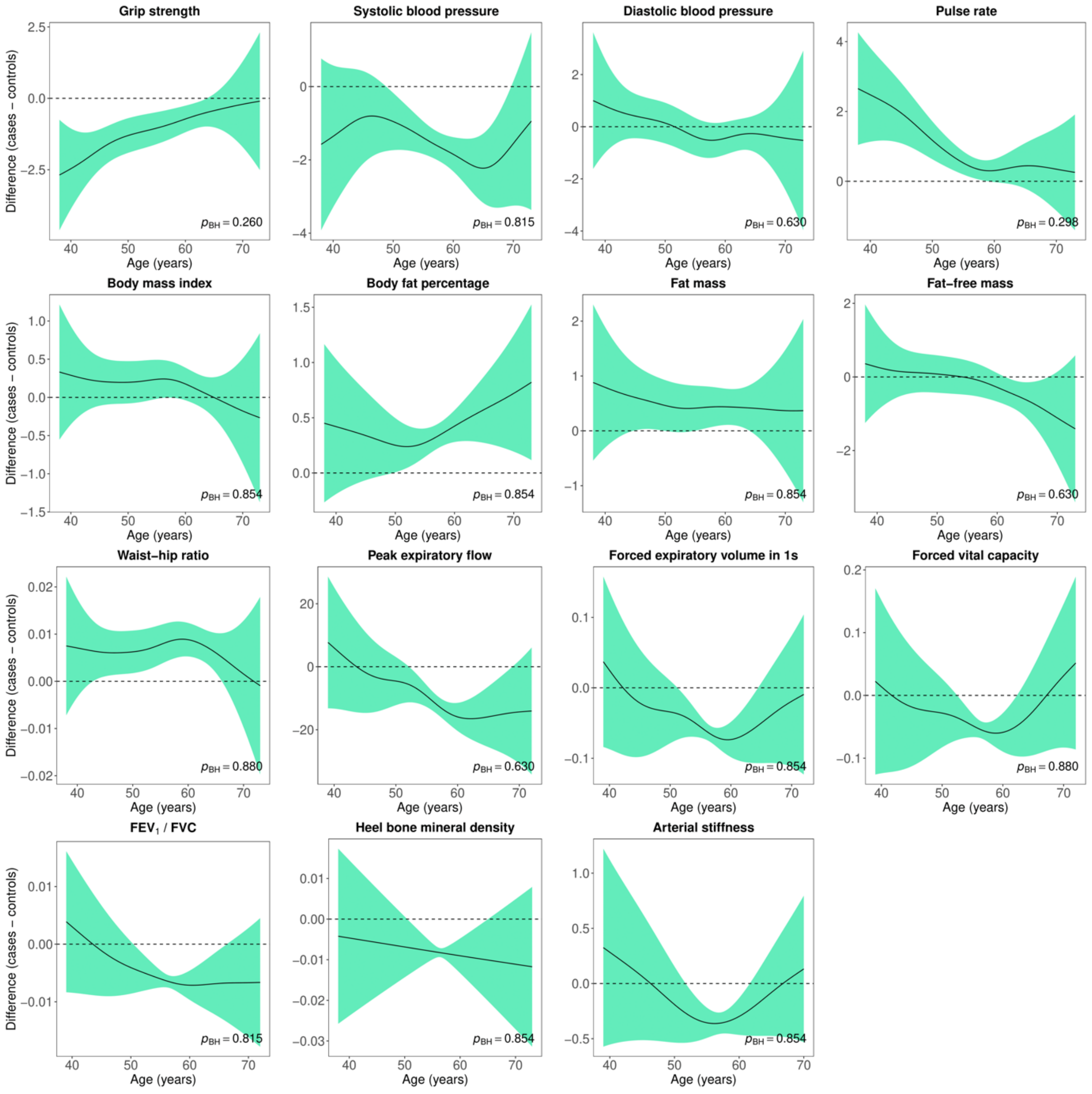

**Supplement figure 11.** Difference smooths comparing age-related changes in physiological measures of males with chronic and/or severe anxiety disorders to healthy controls. Models were adjusted for ethnicity (except lung function), highest educational/professional qualification, physical activity, smoking status, alcohol intake frequency, sleep duration and, for cardiovascular measures, current use of antihypertensive medications. The shaded areas correspond to approximate 95% confidence intervals (± 2 × standard error). Negative values on the y-axes correspond to lower values in males with chronic and/or severe anxiety disorders compared to healthy controls. The horizontal lines represent no difference between female cases and controls. $FEV_1$ = forced expiratory volume in one second; FVC = forced vital capacity; Bonf. = Bonferroni; BH = Benjamini & Hochberg.

**Adjusted GAMs females – anxiety disorders without comorbid depression**

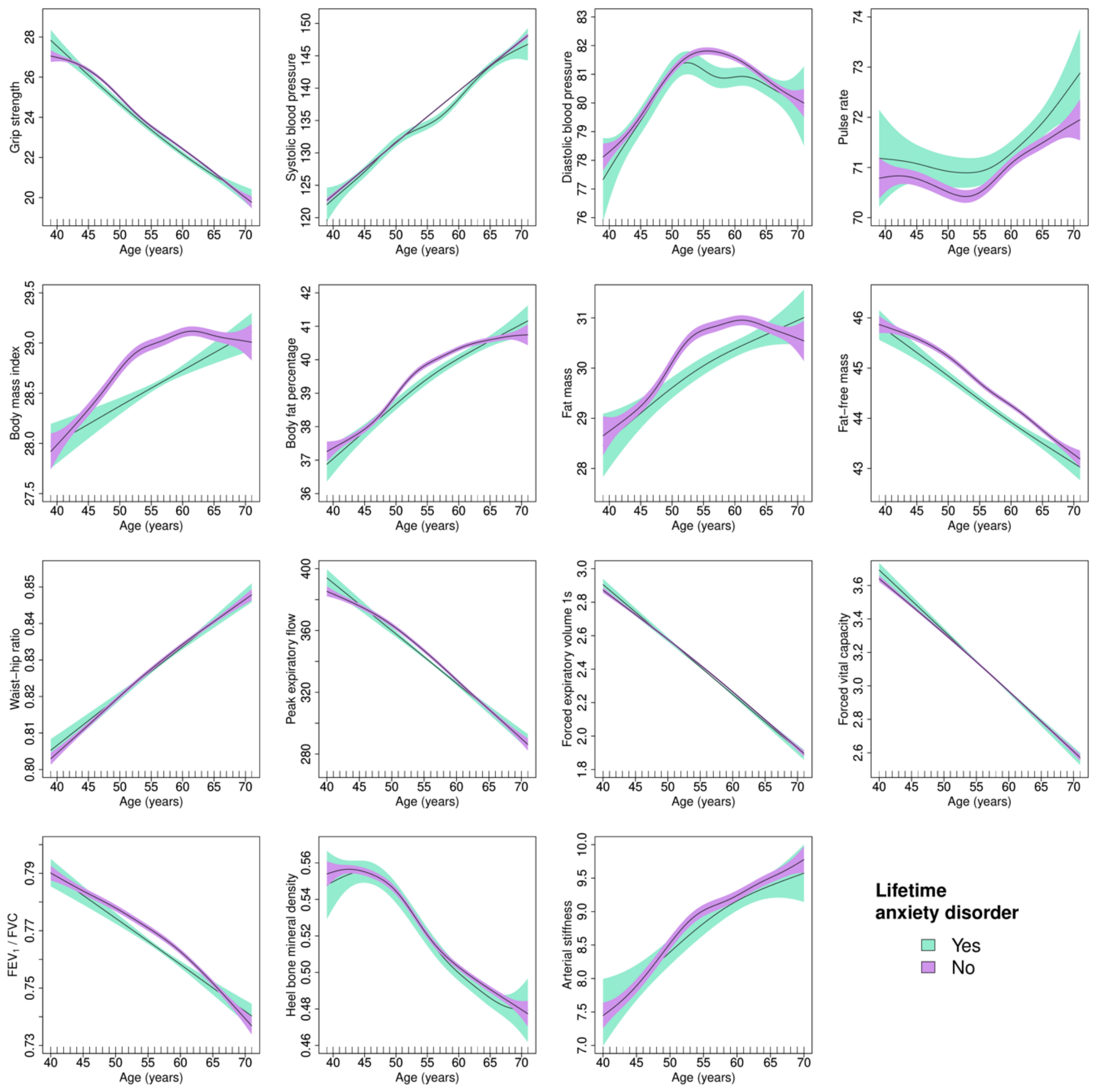

**Supplement figure 12.** Adjusted generalised additive models of age-related changes in physiological measures in females with anxiety disorders without comorbid depression and healthy controls. Models were adjusted for ethnicity (except lung function), highest educational/professional qualification, physical activity, smoking status, alcohol intake frequency, sleep duration and, for cardiovascular measures, current use of antihypertensive medications. The solid lines represent physiological measures against smoothing functions of age. The shaded areas correspond to approximate 95% confidence intervals (± 2 × standard error). $FEV_1$ = forced expiratory volume in one second; FVC = forced vital capacity.

**Adjusted difference smooths females – anxiety disorders without comorbid depression**

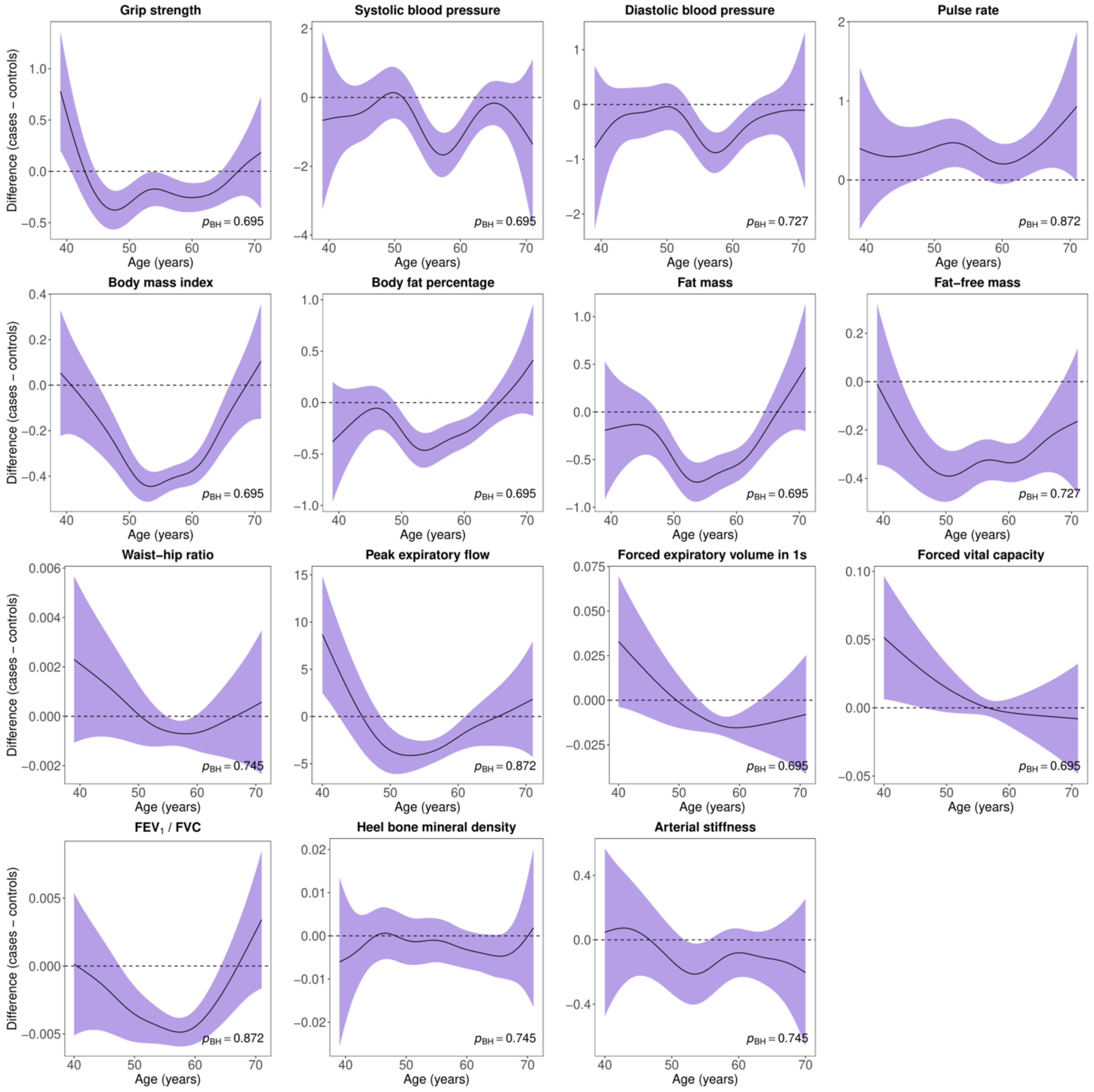

**Supplement figure 13.** Difference smooths comparing age-related changes in physiological measures of females with anxiety disorders without comorbid depression to healthy controls. Models were adjusted for ethnicity (except lung function), highest educational/professional qualification, physical activity, smoking status, alcohol intake frequency, sleep duration and, for cardiovascular measures, current use of antihypertensive medications. The shaded areas correspond to approximate 95% confidence intervals (± 2 × standard error). Negative values on the y-axes correspond to lower values in females with anxiety disorders without comorbid depression compared to healthy controls. The horizontal lines represent no difference between female cases and controls. $FEV_1$ = forced expiratory volume in one second; FVC = forced vital capacity; Bonf. = Bonferroni; BH = Benjamini & Hochberg.

**Adjusted GAMs males – anxiety disorders without comorbid depression**

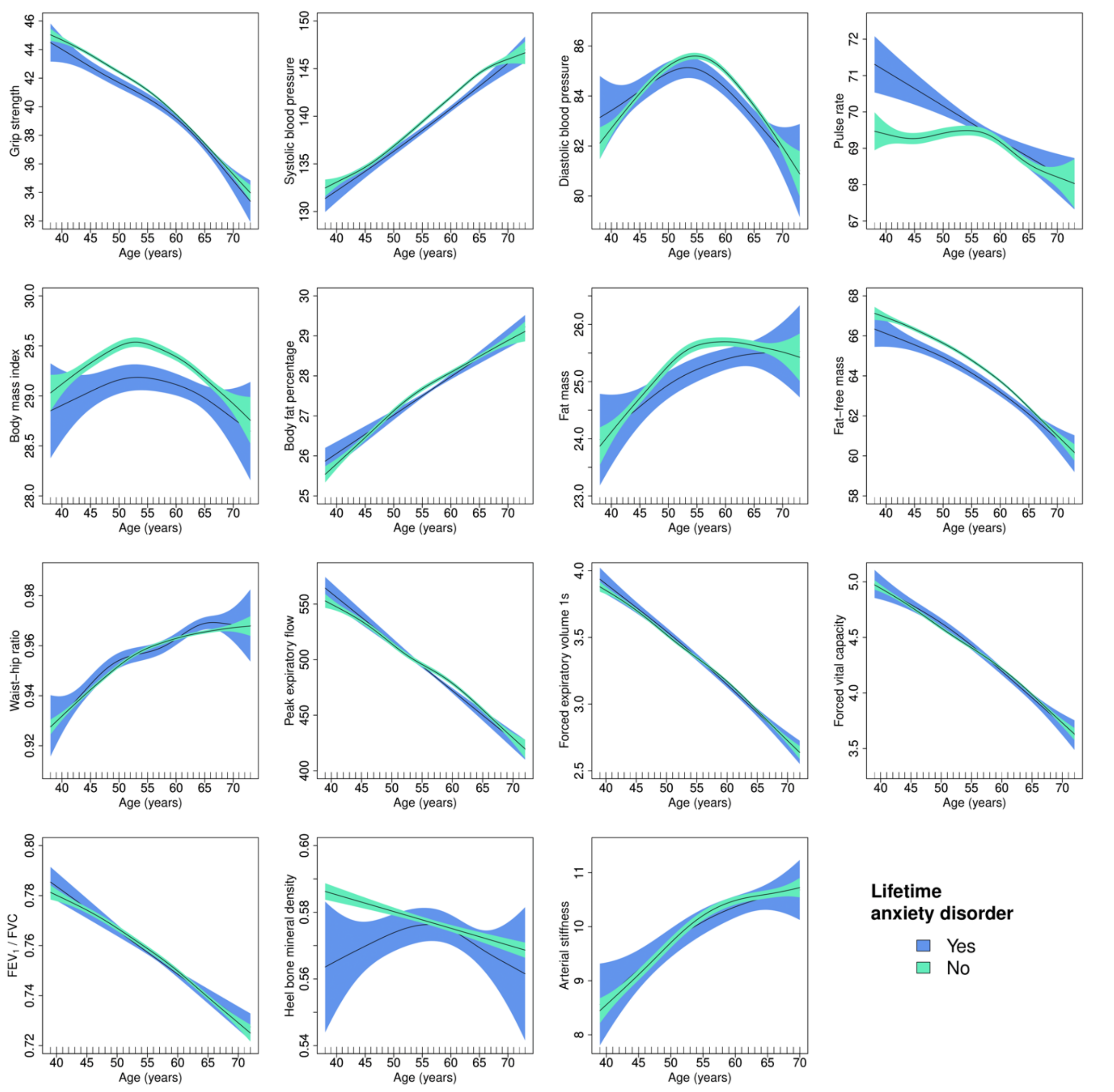

**Supplement figure 14.** Adjusted generalised additive models of age-related changes in physiological measures in males with anxiety disorders without comorbid depression and healthy controls. Models were adjusted for ethnicity (except lung function), highest educational/professional qualification, physical activity, smoking status, alcohol intake frequency, sleep duration and, for cardiovascular measures, current use of antihypertensive medications. The solid lines represent physiological measures against smoothing functions of age. The shaded areas correspond to approximate 95% confidence intervals (± 2 × standard error). $FEV_1$ = forced expiratory volume in one second; FVC = forced vital capacity.

**Adjusted difference smooths males – anxiety disorders without comorbid depression**

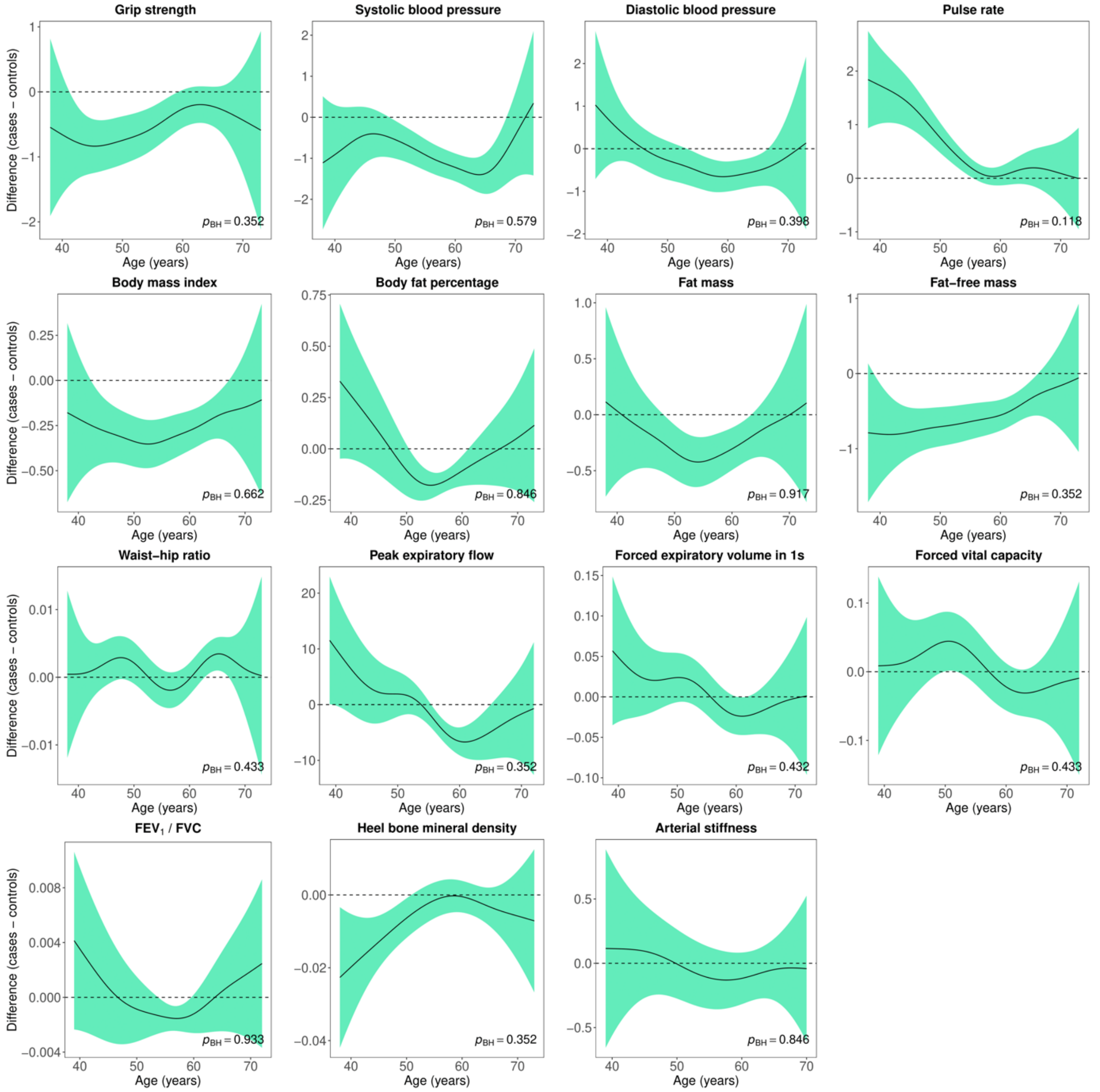

**Supplement figure 15.** Difference smooths comparing age-related changes in physiological measures of males with anxiety disorders without comorbid depression to healthy controls. Models were adjusted for ethnicity (except lung function), highest educational/professional qualification, physical activity, smoking status, alcohol intake frequency, sleep duration and, for cardiovascular measures, current use of antihypertensive medications. The shaded areas correspond to approximate 95% confidence intervals (± 2 × standard error). Negative values on the y-axes correspond to lower values in males with anxiety disorders without comorbid depression compared to healthy controls. The horizontal lines represent no difference between female cases and controls. $FEV_1$ = forced expiratory volume in one second; FVC = forced vital capacity; Bonf. = Bonferroni; BH = Benjamini & Hochberg.

**Adjusted GAMs females – cardiovascular function adjusted for BMI**

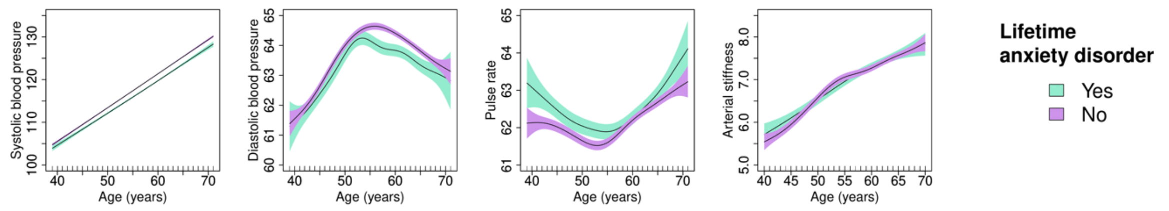

**Supplement figure 16.** Adjusted generalised additive models of age-related changes in cardiovascular function in females with anxiety disorders and healthy controls. Models were adjusted for ethnicity, highest educational/professional qualification, physical activity, smoking status, alcohol intake frequency, sleep duration, current use of antihypertensive medications and body mass index. The solid lines represent physiological measures against smoothing functions of age. The shaded areas correspond to approximate 95% confidence intervals (± 2 × standard error).

**Adjusted GAMs males – cardiovascular function adjusted for BMI**

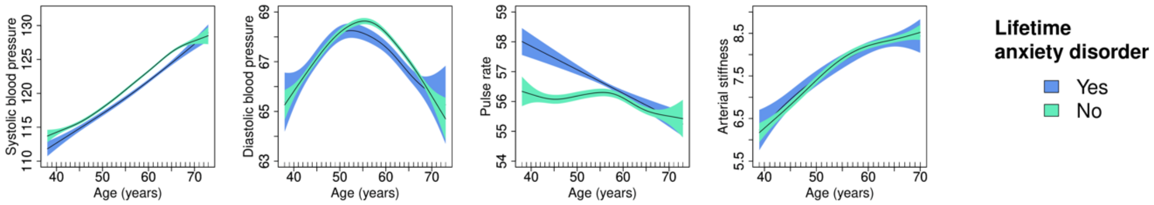

**Supplement figure 17.** Adjusted generalised additive models of age-related changes in cardiovascular function in males with anxiety disorders and healthy controls. Models were adjusted for ethnicity, highest educational/professional qualification, physical activity, smoking status, alcohol intake frequency, sleep duration, current use of antihypertensive medications and body mass index. The solid lines represent physiological measures against smoothing functions of age. The shaded areas correspond to approximate 95% confidence intervals (± 2 × standard error).

### Case-control differences in blood pressure – no antidepressant medication use

**Supplement table 8.** Differences in blood pressure between individuals with anxiety disorders without current antidepressant use and healthy controls

| | Female | | | | | Male | | | | |
|---|---|---|---|---|---|---|---|---|---|---|
| Variable | SMD | 95% CI | | $p_{Bonf.}$ | $p_{BH}$ | SMD | 95% CI | | $p_{Bonf.}$ | $p_{BH}$ |
| Systolic blood pressure | -0.119 | -0.133 | -0.105 | <0.001 | <0.001 | -0.085 | -0.103 | -0.067 | <0.001 | <0.001 |
| Diastolic blood pressure | -0.049 | -0.063 | -0.036 | <0.001 | <0.001 | -0.035 | -0.054 | -0.017 | 0.002 | <0.001 |

*Note:* SMD = standardised mean difference; CI = confidence interval; Bonf. = Bonferroni; BH = Benjamini & Hochberg. *P*-values for Welch's t-test. Negative values correspond to lower values in anxiety disorder cases.
